# Baseline nowcasting methods for handling delays in epidemiological data

**DOI:** 10.1101/2025.08.14.25333653

**Authors:** Kaitlyn E. Johnson, Maria L. Tang, Emily Tyszka, Laura Jones, Barbora Nemcova, Daniel Wolffram, Rosa Ergas, Nicholas G. Reich, Sebastian Funk, Jonathon Mellor, Johannes Bracher, Sam Abbott

## Abstract

**Background:** Up-to-date real-time disease surveillance data can provide critical public health insights, however reporting delays can create downward bias in the latest data. Nowcasting methods designed to correct for this bias remain underused in public health practice due to their complexity, lack of tailored documentation, or technical barriers. Methodological advances in nowcasting are also hampered by the absence of standardised benchmarks for evaluating new methods.

**Methods:** To address these needs, we developed a family of nowcasting methods and an accompanying R package, *baselinenowcast*. We validated our method against the baseline method that was used in the German COVID-19 Nowcast Hub and on which our approach was based. Using this data, we conducted an analysis to compare different specifications of our method which were designed to address common issues in epidemiology such as weekday patterns in reporting and the ability to share estimates across different strata. We used our approach on norovirus surveillance data from the United Kingdom Health Security Agency (UKHSA) and compared the performance of three of our method specifications against three methods evaluated in a previous study.

**Results:** Our baseline method improved estimates compared to unadjusted data across all case studies. We found that the optimal choice of baseline method specification depends on context but that our default method specification performed well in a range of settings. Applied to UKHSA norovirus data, our method helped us understand the performance of the model currently used in public health practice.

**Conclusions:** Our method and software can be used both as a straightforward nowcasting method and provides a benchmark for nowcasting model development.

**Plain Language Summary:** Reporting delays in public health surveillance systems can create a misleading impression of declining trends in recent data. While a number of “nowcasting” methods have been developed to correct for this bias, widespread adoption in public health practice has yet to be realized. Currently, there is no simple method to perform nowcasting that both meets the needs of public health practice and can be used as a benchmark for further methodological advancement. To address these two gaps, we developed a family of baseline nowcasting methods and an accompanying software package. Using data from COVID-19 in Germany and norovirus in England, we evaluated the performance of our default method against other methods used in previous studies and assessed the performance of our different method specifications designed specifically for common problems in epidemiology such as weekday patterns in reporting and sharing estimates across different strata. Our findings indicate that our baseline methods improve performance over unadjusted data or more ad-hoc baseline nowcasting approaches, provide an interpretable and accessible nowcasting solution for public health practice, and are useful as a standard benchmark for enhanced understanding of more advanced methods.

## Introduction

Delays in epidemiological data present a challenge in public health surveillance, as they can create a misleading impression of declining trends in recent data. This bias is due to “right-truncation” of primary events (e.g. symptom onset or hospital admission), where recent events are systematically under-represented because reporting delays cause some portion of these events to not yet be present in the real-time surveillance data. This affects numerous surveillance systems across pathogens, including COVID-19, mpox, and norovirus [1–7]. Recently, there have been several applications of nowcasting methods aimed at correcting for this downward bias. Examples include public facing data dashboards such as at Massachusetts Department of Public Health [8], Dutch National Institute for Public Health and the Environment (RIVM) [9], and the U.S. Centers for Disease Control and Prevention (U.S. CDC) [10], and internally at UK Health Security Agency (UKHSA) for norovirus, flu, and RSV nowcasting, Robert Koch Institute (RKI) for acute respiratory incidence nowcasting, and the U.S. CDC for estimation of the effective reproductive number for flu and COVID-19, on publicly available data [3,11,12], and as part of outbreak responses [13,14]. However, these applications still remain the exception, with the majority of potential use-cases handled by showing uncorrected data or shortening time series to remove partially complete data, which has the effect of reducing the amount of information available on the latest trends [2,15].

Several challenges remain in addressing systematic downward bias in surveillance systems due to right-truncation of primary epidemiological events in public health practice. Frontline practitioners require methods that are statistically rigorous, straightforward to implement, and can be easily communicated to stakeholders. However, no currently available tools meet these needs fully. The chain ladder method [16] is an empirically-driven approach that could meet the needs of practitioners as it is close to the kinds of methods often implemented in real world settings. However, current implementations of the method either lack domain-specific features and documentation (e.g. the *chainladder* R package [17] designed for nowcasting insurance claims) or have been implemented in an ad-hoc manner rather than intended for general applicability [3,8]. Other methods designed specifically for nowcasting epidemiological data are often complex due to the flexibility and feature set that supporting a range of epidemiological settings requires [18–22], leading to a high barrier to entry. This means practitioners may have to choose between delaying insights (by not making full use of recent data), risk misleading interpretations from unadjusted data, developing methods that may lack reusability across different contexts or not be fully validated, or use methods not appropriate for their context. For methodological research, the absence of standardised benchmarks makes understanding the value of novel methods more challenging. In other fields and other settings, baseline methods, which are relatively straightforward methods to implement and serve as a reference point for evaluating other methods [23], have been suggested as a potential solution for this issue [24]. Without consistent baselines for evaluation, researchers and public health practitioners may struggle to quantify incremental improvements or understand trade-offs between model complexity and performance across different epidemiological contexts [23]. The chain ladder approach has been used on an ad-hoc basis to meet this need in [3,8,25]. For wider usage, a standardised method is needed.

We propose a family of baseline nowcasting methods and an accompanying R package, *baselinenowcast*, designed to address these dual needs in infectious disease surveillance, building on the empirically-driven chain ladder implementation described by Wolffram et al. [3,16,17]. In this work, we validate our approach by comparing the Wolffram et al. [3] method against our baseline method with a matching specification using German COVID-19 data. We then compare the performance of our different method specifications applied to both the German COVID-19 data and UKHSA norovirus surveillance data [2] alongside existing nowcasting methods.

## Methods

### Input data requirements: reporting triangle

Our method requires the number of incident cases indexed by the reference time (e.g. date of symptom onset, specimen collection, or hospital admission) and report time (e.g. the date a positive test is reported into a surveillance system). Data in this format allows us to reconstruct reporting delays (i.e. the time from the reference time to the report time). Combining these reporting delays results in a reporting delay distribution (i.e the distribution of times from the reference time to the time that a case is reported) for each reference time. In wide format, where each reference time is a row and each reporting delay is a column, this data is known as the “reporting triangle” [26] because the observations form a triangle with the bottom right elements not yet observed as of the most recent reference time. The goal of nowcasting is to estimate these unobserved events. If we do not do so, and instead sum only the observed columns, then our case counts will be systematically downward biased. The fully observed version of these data, the “complete reporting matrix”, can be used to estimate the reporting delay distribution as each row can be normalised by the sum of that row to give the proportion reported at a given reporting delay. It is data of this format, or reconstructions thereof, that we use in our nowcasting method.

### Statistical framework

#### Overview

Our approach, based on the chain ladder method in the version described by Wolffram et al. [3], consists of five key steps, each of which are separate modular operations that can be modified (Figure 2). In the following, we give a high-level overview of the “default” specification of our method, its data ingestion settings and supported variations. For the full details see the SI Text. *Mathematical model*.

**Figure 1.**
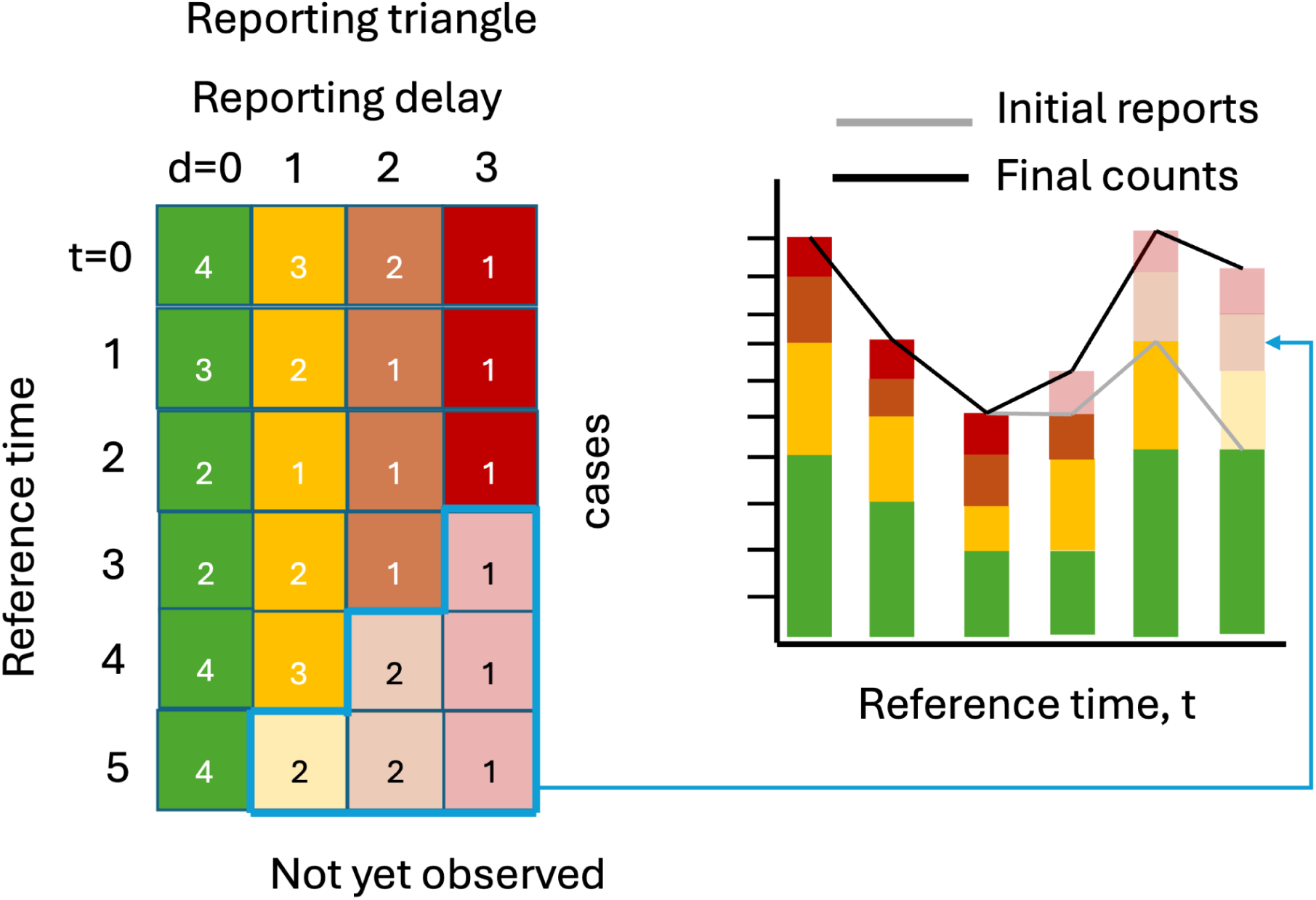
Illustration of the reporting triangle and corresponding case counts aggregated by reference time. Rows of the reporting triangle represent reference times and columns represent reporting delays with numbers indicating the incident case count at that reference time and reporting delay. Colors indicate the delay, with solid shading indicating observed case counts and transparent shading indicating not yet observed cases, as of the most recent reference time. The bar chart shows the cases summed by reference time, with gray lines indicating the total number of cases reported at each reference time as of the most recent reference time, and black lines indicating the eventually observed number of cases. Figure 1 depicts a visual representation of the reporting triangle on the left and the corresponding case counts aggregated by reference time on the right. The solid shading indicates what has been observed as of the most recent reference time, and the lighter shading indicates what will eventually be observed. If we were to aggregate cases by reference time as of the most recent reference time, we would be missing cases from delays 1, 2, and 3 for the most recent reference time, delays 2 and 3 from the second most recent reference time, etc., resulting in systematic downward bias in the recent trend, as indicated by the gray line in the bar chart. Eventually, the counts would be revised upwards as the elements of the reporting triangle are observed at later delays, resulting in the trend in the final counts being corrected (black lines).

**Figure 2.**
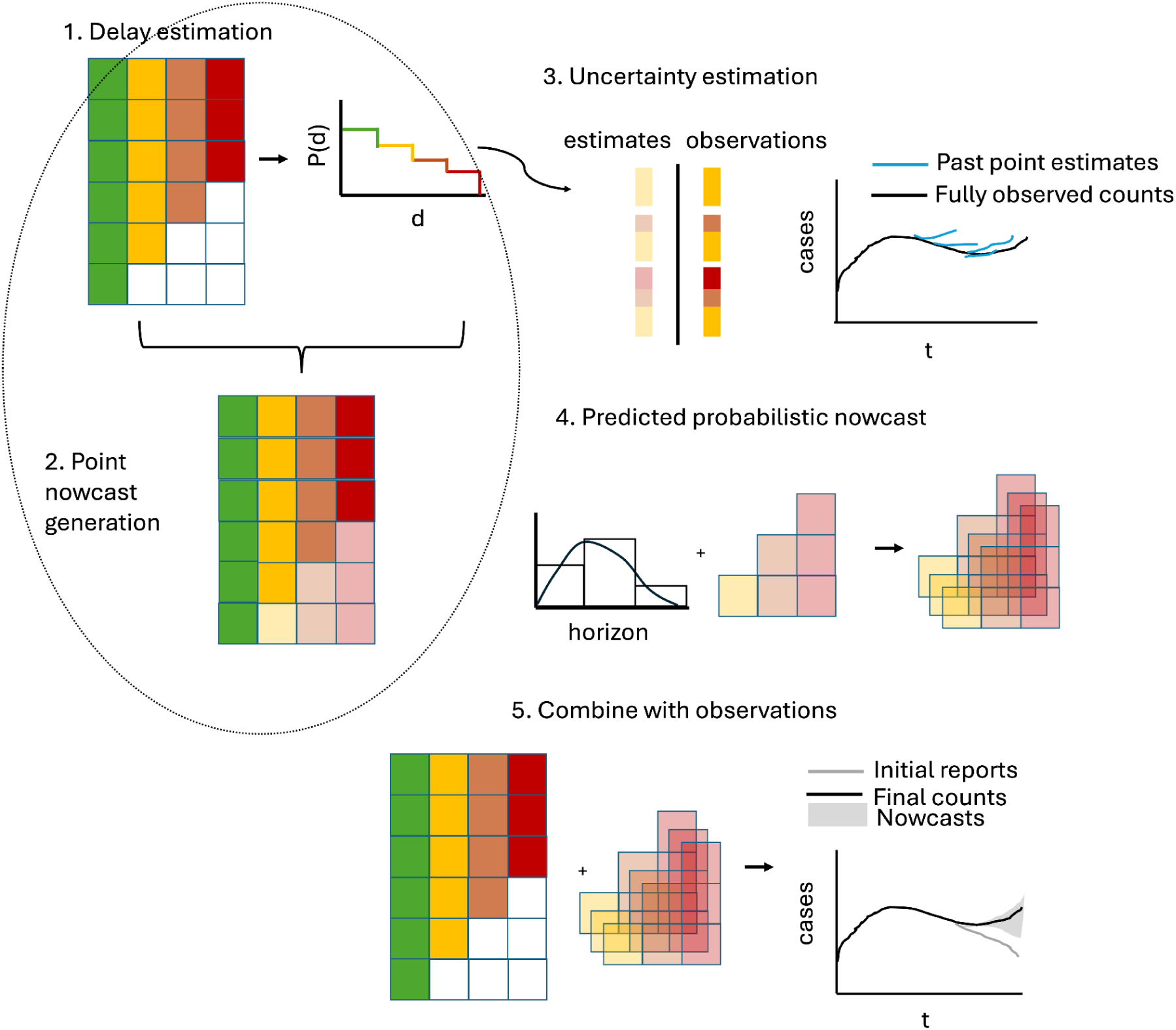
Schematic illustration of the *baselinenowcast* method. The reporting triangle contains the number of cases by reference time (rows) and reporting delay (columns), where colors represent the delays, and with white indicating observations that have not yet been observed and shading here indicating whether values have been observed (solid) or imputed (shaded). The delay distribution is estimated empirically from the reporting triangle. Point nowcasts are generated using the delay distribution and the partial observations at each reference time to impute the missing values in the bottom right of the triangle. Uncertainty is quantified by repeating this procedure for a number of past retrospective nowcast times and comparing the estimates to the observations. The point nowcast predictions and the uncertainty parameters are used to generate probabilistic predictions, and these are combined with the observations to generate probabilistic nowcasts.

##### Step 1: Delay estimation

- The empirical delay distribution can be estimated directly from a complete reporting matrix by dividing the sum of each column by the total count.
- By default we use the partial observations from the most recent data points in order to incorporate the most up-to-date data on the delay distribution. To do so, we need to first “fill in” the missing observations in the reporting triangle (white boxes in bottom right of reporting triangle, Figure 2) by assuming that the delay distribution is constant over time and:

a. First compute the ratio between the first delay and the second delay for all reference times when both delays are fully observed (e.g. the ratio of the sum of the first 5 green vs yellow rows in Figure 1 (4+2+2+3+4)/(3+2+1+2+3) = 1.36)
b. Divide the partially observed row of the first delay column by the ratio to impute the missing component of the second delay (e.g. the case count at reference time = 5 and delay = 0, 4, from Figure 1, is used to impute the case count at reference time = 5, delay = 1 as 4/1.36 = 2.93)
c. Repeat for the missing elements of each column filling in from left to right.

##### Step 2: Point nowcast generation

- Given a delay distribution and a reporting triangle, we impute the missing observations to generate a point nowcast matrix: a complete reporting matrix where the missing observations have been filled in with point estimates (indicated by the shaded bottom right of the triangle in Figure 2, step 2). The rows sum to a point estimate of the final counts at each reference time.
- We use a similar approach to that used to impute the missing elements of the reporting triangle in step 1 but now we:

a. Estimate the final case counts by dividing the current total reported counts by the proportion estimated to be reported by this time (e.g. if there are 4 case counts observed so far at a reference time of 5 and a delay of 0 from Figure 1, which is estimated to represent 40% of eventual cases from the delay distribution in step 1 of Figure 2, we expect 4 / 0.40 = 10 total final cases)
b. For each unobserved delay, we multiply the estimated final case counts by the proportion of cases observed on that delay (e.g. if an estimated 10% of cases are reported with a delay of 3 from the delay distribution in step 1 of Figure 2 and we expect 10 total cases, we expect 10 * 0.1 = 1 to be reported with a delay of 3)
- We implement a slightly more complex version of this to account for zero reported cases observed so far.

##### Step 3: Uncertainty estimation

- We use past nowcast errors compared to observed values as of the current nowcast date to quantify uncertainty.
- We generate retrospective point nowcasts (light shaded bars and blue lines in Figure 2, step 3) by repeating steps 1 and 2 for a number of past, retrospective nowcast times.
- An observation model, which is by default a negative binomial, is fit to the predicted (shaded bars, blue lines Figure 2, step 3) and observed (solid bars, black lines Figure 2, step 3) components for each nowcast horizon.

##### Step 4: Predicted probabilistic nowcast

- We generate probabilistic predictions by sampling draws (indicated by the stacked shaded bottom right reporting triangles in Figure 2, step 4) from the observation model at each horizon with a mean given by the point nowcast and the horizon specific uncertainty parameter (bar chart with distributional overlay in Figure 2, step 4) estimated in step 3.
- By default, the target we compare to is the sum of the not-yet observed components of the point nowcast for each reference time.

##### Step 5: Combine with observations

- We then add what has already been observed (reporting triangle Figure 2, step 5) to each draw (shaded bottom right of reporting triangle Figure 2, step 5) of the predicted probabilistic nowcast for what has yet to be observed to create a complete probabilistic nowcast of the final counts for each reference time point (indicated by gray shading in line plot Figure 2, step 5).

As well as model choices, we also make data ingestion assumptions in our default approach. These are all conditioned on the maximum delay observed in the data making them more robust to different datasets. The maximum delay is set by the user when creating the reporting triangle based on when the data is expected to stabilize, with shorter maximum delays being more computationally efficient.

1. **Overall data use:** We use three times the maximum delay number of reference times (in line with Wolffram et al. [3]).
2. **Delay distribution estimation:** 50% of the reference times are used for delay estimation, with a minimum requirement of at least one more than the number of reference times with partial observations.
3. **Uncertainty Estimation:** 50% of the reference times are used as retrospective point nowcast times. If there is insufficient data, the method uses the oldest retrospective nowcast time with at least the same amount of historical data for delay estimation as is used for the point nowcast at the current nowcast time, and requires that there are at least two retrospective nowcasts containing sufficient data.

#### Available options for model specification

As our method is modular with each step being independent, there are a large number of potential variations, designed with common scenarios in infectious disease surveillance in mind. We have implemented the following:

##### Input data options

- Use of a complete reporting matrix or a reporting triangle.
- Changing the total number of reference time points used to estimate the delay distribution and uncertainty parameters, balancing precision (from more data) against capturing recent variability (from more current data).
- Reference and report dates of different temporal granularities, e.g. daily reporting of weekly reference dates, weekly reporting of daily reference dates, and weekly reference and report dates.

##### Delay estimation approaches

- Estimate delays from the same data set / stratum being nowcasted or “borrow” estimates from another reporting triangle. This may be useful for sparse data sets, such as in small subpopulations or locations or for rare pathogens and emerging outbreaks.
- Changing the proportion of data used to estimate the delay with the same tradeoffs as for changing the overall amount of data input.
- Adjusting the maximum delay, but this should be based on empirically observed delay rather than being viewed as a modelling option.

##### Uncertainty quantification variations

- Observation models can be fit using different datasets than those used for delay estimation or for the nowcast.
- Changing the proportion of data used to estimate the observation model with the same tradeoffs as in the delay estimation case.
- The parametric form of the observation model can be changed.
- The observation model can be fit jointly for any grouping of observations. The target choice for the final nowcast can be specified such that the uncertainty estimation occurs on this target (i.e. 7-day rolling sum).

#### Software implementation

Our baseline nowcasting methods are implemented in the R package *baselinenowcast*, which provides a modular pipe-friendly interface for nowcasting right-truncated surveillance. The package has minimal dependencies, primarily relying on base R functionality [27]. A primary workflow function *baselinenowcast()* implements the default method, whilst individual component functions enable customisation of specific steps through a consistent interface. The package accepts input data in a standard data frame format with columns for reference date, report date, and count. An additional feature is its compatibility with the *epinowcast* R package objects [19], allowing users to leverage that package’s preprocessing and data conversion functions and facilitating comparison to more *epinowcast* nowcasting models. Low-level functions use a matrix input for efficiency. We followed software best practices, including unit testing, continuous integration, code review via pull requests, and documentation driven development throughout the package’s development (https://github.com/epinowcast/baselinenowcast) [28,29].

### Evaluation analysis

To evaluate nowcast performance from quantile-based outputs, we use the weighted interval score (WIS). WIS is a widely used proper scoring rule used to assess forecast quality of probabilistic quantile-based forecasts [23,30,31] and is the probabilistic equivalent of the absolute error of point forecasts. We decompose WIS into overprediction, underprediction, and dispersion components. As WIS is an absolute measure of error, we compute the relative WIS throughout, with the default specification of our baseline method used as a comparison unless otherwise noted. We assess forecast calibration using coverage metrics, where we compute the proportion of all observations within a specified prediction interval. For example, a well-calibrated forecast would have 50% of the observations fall within the 50% prediction intervals (between the 25th and 75th quantiles). See the following sections for details on the quantiles used to compute the WIS scores and coverage metrics.

#### German COVID-19 Nowcast Hub validation

To verify that our *baselinenowcast* implementation worked as expected and to characterize the performance of our method overall and across age groups and nowcast dates, we produced nowcasts using our method’s default specification and compared them to the nowcasts produced using the the baseline nowcasting method described in Wolffram et al. [3] (referred to in that paper as KIT simple nowcast) which our method and default model settings were based upon. Both methods were applied to the COVID-19 hospitalization data from the German COVID-19 Nowcast Hub (the Hub) [32]. The dataset represents 7-day rolling sums of hospitalisation incidences due to COVID-19 in Germany, which were used to guide public health policy. German hospitalization counts were aggregated by the date of positive test result rather than admission (see Wolffram et al. [3] for details). They were hence particularly affected by delays, as these not only comprised the time from hospitalization to report, but also from positive test to hospitalization. Data then sometimes took several weeks or months to fully stabilize. We used all the available data spanning from 22 November 2021 to 29 April 2022, with preliminary data available from 1 July 2021. We used the pre-processed dataset available in the Hub’s GitHub repository [32], which contains a cleaned version of the reporting triangle that teams submitting to the Hub were instructed to use.

We generated nowcasts of the 7-day rolling sum of COVID-19 hospitalisation incidence in Germany each day for national-level aggregated data and age-stratified data. We applied our baseline nowcast with a maximum delay of 40 days, consistent with the specification of the KIT simple nowcast model used in [3], and with a training volume of 120 reference dates used by each nowcast (this means that 60 reference dates were used for delay estimation and 60 retrospective nowcast datasets were created for uncertainty estimation). For consistency with the KIT simple nowcast model, we estimated uncertainty in the predicted components of the final “target”, which in this case was the 7-day rolling sum of the partial expectation of hospitalisation incidence. This made our baseline approach theoretically equivalent to the KIT simple nowcast model described in [3], except that they did not include a correction for zero counts in the partial observations for a given reference time.

However, when verifying our method against the KIT simple nowcast codebase, we identified an issue impacting the dispersion estimate (see SI Text. *Description of KIT simple nowcast implementation issue and revised nowcasts* for more details). We fixed the issue in our fork of the repository and retrospectively generated new nowcasts, which we labeled as the “revised KIT simple nowcast”. We use this as a point of comparison in the main text results, with a comparison to the original implementation detailed in the supplement and referred to as the “original KIT simple nowcast”.

We visualised summary statistics derived from the data. We plotted the mean reporting delay over time for national-level aggregations and individual age groups to identify any temporal patterns. We also visualised the full distribution of delays nationally and by age group to characterise differences in reporting processes across strata.

We then visualised the nowcasts against the eventually observed values to qualitatively verify equivalence for a subset of nowcast dates. We then quantitatively evaluated both sets of nowcasts using a rolling evaluation dataset of 40 days after each nowcast date, restricted to a maximum delay of 40 days for each reference date, consistent with the analysis performed in section 3.7 of Wolffram et al. [3]. This means that for each nowcast date, we used a different evaluation dataset corresponding to the data available 40 days after that nowcast date. We evaluated nowcasts for horizon days 0 to - 28 days. We used the same quantiles as in [3] (0.025, 0.1, 0.25, 0.5, 0.75, 0.9, 0.975) to compute the WIS. We computed WIS scores for our baseline method, the revised KIT simple nowcast, and the original KIT simple nowcast. We also calculated the empirical coverage at the 50% and 95% prediction intervals. We calculated the relative WIS across multiple dimensions: by nowcast horizon (from 0 to - 28 days), by nowcast date, and by age group.

#### Performance of different *baselinenowcast* method specifications applied to German COVID-19 data

To assess whether other possible *baselinenowcast* method specifications designed specifically for the epidemiological context could further improve performance, we performed a sensitivity analysis of the following pairwise method comparisons: borrowing delay and uncertainty estimates from national-level data when nowcasting for specific age groups, using only complete reporting matrices versus reporting triangles, using shorter (50%) and longer (200%) training periods than those used in the default specification, and estimating delays independently for each weekday and combining.

We generated nowcasts of the 7-day rolling sum of COVID-19 hospitalisation incidence in Germany each day for the same period as in the validation study for all age groups with each method specification. We visualised predictions against the 40-day ahead rolling evaluation dataset values, focusing on same-day nowcasts, and again used the weighted interval score (WIS) and coverage metrics for quantitative assessment, using the same quantiles and prediction intervals as described in the validation section. We again stratified our analysis by nowcast horizon (from 0 to - 28 days), and by age group. We calculated WIS for all method specifications and relative WIS compared to the default method specification.

#### Case study: UKHSA norovirus surveillance

To further validate our approach and explore its utility as a baseline method we produced nowcasts using three specifications of our baseline method for UKHSA’s norovirus surveillance data, and compared the performance of those nowcasts to the performance of three models in Mellor et al. [2] including an alternative baseline approach. These data consist of laboratory-confirmed norovirus cases in England from winter 2023/2024. Each case record includes a specimen date (when the sample was collected) and a report date (when the test result was ingested into the surveillance system). The difference between these dates forms the reporting delay, which typically peaks at 1-2 days with most reports occurring within 7 days. However, approximately 20% of specimens obtained in a given week were not reported by the end of that week, creating a substantial right-truncation effect in recent data. Because of the variable results across days of the week observed in the original study, we assumed that the dataset exhibited strong day-of-week-periodicity, with reporting patterns varying systematically across weekdays.

We implemented our baseline nowcasting method using three different method specifications. All method specifications used a maximum delay of 14 days to match the approach in the original study for the Generalized Additive Model (GAM) model. The first method was our default baseline method, except that we fit to the most recent 56 days of data rather than the default of 42 days (3 times the maximum delay of 14 days). This was motivated by following the settings used for the GAM model so that the methods used equivalent amounts of training data and so were more directly comparable. We refer to this method specification as the base specification. Based on our hypothesis that day of the week variations in the delays were present we used two further methods. The second method independently estimates nowcasts for each day of the week, using the most recent 11 weeks of data (11 data points per day of week), to use a similar amount of data to the base specification (56 days), though in practice we needed more than 56 days of data due to data sparsity in some weekdays. The third model had the same structure as the second model but with more training data, instead using the 56 latest data points from each particular day of the week (spanning approximately 56 weeks of historical data) or as much as was available, to use the same amount of training data as the base specification for each weekday’s delay estimation. We refer to these method specifications as the *baselinenowcast* weekday filter small/large training volume specifications respectively.

Following Mellor et al. [2], we nowcast each Sunday from November 5th 2023, to March 10 2024 and evaluated performance on horizons 0 to -7 days using rolling 50-day ahead of the nowcast date evaluation datasets (similar to the rolling evaluation applied to the German COVID-19 data). For consistency with the evaluation in Mellor et al. [2], we did not restrict the evaluation dataset to only reports within the maximum delay. We then compared nowcast performance from three of the models considered in Mellor et al. [2]: a GAM, an implementation of epinowcast, and a naive baseline method. The GAM was developed by the authors of the original study [2] and uses a latent model of the number of cases as a function of reference time and delay, accounting for weekday effects in the reference time and the reporting delay. The epinowcast implementation uses the R package *epinowcast* [19] to estimate a Bayesian hierarchical model of the number of observed cases by reference time and delay, using a latent variable approach to model the underlying trend in the epidemic growth rate accounting for weekday effects in the underlying infections. The Mellor et al. [2] baseline implementation assumes that the cases predicted each day of this week will be equal to the observed count from the previous week (essentially propagates the previous week forward), producing only a median nowcast.

We first visualised the nowcasts against the eventually observed values to qualitatively assess performance. To align with Mellor et al. [2], we calculated WIS using only the 50% and 90% prediction intervals. Coverage metrics were also calculated using the 50% and 90% prediction intervals. We otherwise followed the evaluation approach of our earlier case studies, using our base specification as the baseline for the relative WIS calculation. We stratified by day of week in addition to week of nowcast, and stratified empirical delays by day of week as well as over time. Unlike the original paper, which examined both daily and weekly aggregated scores, we focused on daily scores.

#### Evaluation analysis implementation

Our analysis pipeline was implemented using *targets* [33] for reproducible workflow management, with *scoringutils* [31,34] used for nowcast evaluation. All analyses were performed in R [27] with key dependencies including *dplyr* [35] for data manipulation and *ggplot2* [36] for visualisation. The code and analyses are available at https://githhub.com/epinowcast/baselinenowcast-paper [37,38].

## Results

### German COVID-19 Nowcast Hub validation

Because our baseline method’s implementation is a generalization of the KIT simple nowcast used by Wolffram et al. [3], we initially verified our method by comparing our baseline method’s results against retrospective nowcasts generated directly from the KIT simple nowcast codebase using the same input data. In this process, we identified a difference in the outputs despite following identical model specifications and having nearly identical mathematical methods, which led us to further investigate the method implemented in the original KIT simple nowcast codebase. We then identified a bug in the original implementation (see SI Text. *Description of KIT simple nowcast implementation issue and revised nowcasts* for more details). We consulted with the authors, whom are co-authors of this work, fixed the bug in our fork of the repository, and regenerated retrospective nowcasts which we have labeled as the revised KIT simple nowcast, with comparisons to the original implementation in the Supplement labeled as the original KIT simple nowcast (SI Figure 1).

Our method produced equivalent nowcasts to the revised KIT simple nowcast (Figure 3A, 3B, 3C). Compared to the original KIT simple nowcast, our baseline method performed better, with an overall relative WIS of 0.88 nationally and 0.91 by age groups (SI Figure 1A, B), indicating that the bug fix improved nowcast performance. Exploratory data analysis indicated that the delay distribution (Figure 3E) differed between age groups, with the 00-04 age group having the shortest mean delay of 3.71 days whilst the 35-59 age group experienced the longest delays, averaging 5.88 days. The mean reporting delay showed changes over time roughly on the time scale of changes in case counts (Figure 3E, 3A).

**Figure 3:**
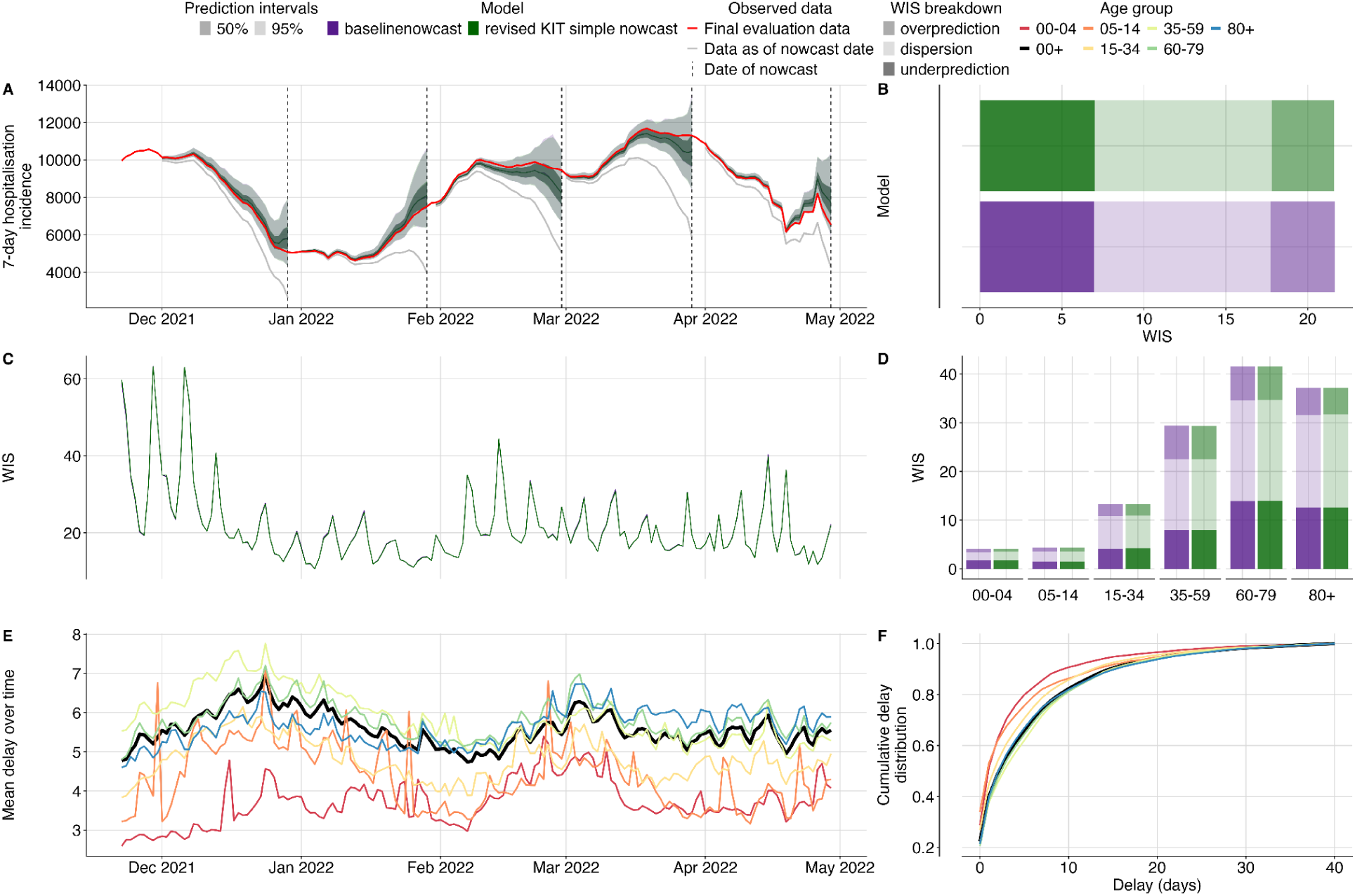
Validation of our baseline nowcasting method using German COVID-19 data compared to the revised KIT simple nowcast model. A. Nowcasts from our baseline model and the revised KIT simple nowcast model for all age groups plotted against eventually observed values aggregated across all age groups, coloured by model. Because both sets of nowcasts were nearly identical, coloring and shading is not distinguishable between the two. Observed data are shown as solid lines with gray indicating data available at the nowcast date and red indicating the final evaluation data. Dashed vertical lines indicate the date of the nowcasts. Shading indicates prediction intervals of each nowcast. Nowcasts are shown from horizons 0 to -28 days. B. Decomposed overall WIS aggregated across age strata. Shading indicates WIS component (dispersion, overprediction, underprediction), colour indicates model. C. Daily mean WIS scores summarised across age strata and 28 horizon days, coloured by model. Identical WIS scores over time renders the two lines indistinguishable. D. Decomposed WIS scores across age groups. Shading indicates WIS component, colour indicates model. E. Mean reporting delay over time. Colour indicates age group, with the national average highlighted in black. Mean reporting delays were calculated using the last 40 reference dates, corresponding to the maximum delay, for each nowcast date and age group. F. Cumulative distribution functions (CDFs) of the mean delay distribution across the entire study period. Colour indicates age group with the national average highlighted in black.

Our baseline method was consistently less biased than no adjustment (gray line Figure 3A). Underprediction, overprediction, and dispersion contributed approximately 32%, 18%, and 50% respectively of the total score across age groups. Our method had 55% coverage at 50% prediction intervals and 96% coverage at 95% prediction intervals, indicating it had slightly wider prediction intervals than observed in the data (SI Figure 6,7). There was moderate variation across nowcast dates that appeared to be correlated with changes in reporting delays and peaks in case counts (Figure 3A, 3C, 3E), as is to be expected as higher case counts will be associated with scores of a higher magnitude. Across age groups, WIS ranged from 4.05 for the 00-04 age group to 45.76 for the 60-79 age-group, again reflecting the magnitude in case counts with 00-04 contributing the fewest hospital admissions and 60-79 year olds contributing the most admissions (Figure 3D).

### Performance of different *baselinenowcast* model specifications applied to German COVID-19 data

We found varying patterns of relative performance compared to the default configuration, depending on age group and method specification (Figure 3). Borrowing delay and uncertainty estimates for each age group from aggregate data reduced nowcast performance (relative WIS (rWIS) of 1.14, Figure 4B). Using only the complete reporting matrices resulted in a slight reduction in performance overall (rWIS 1.04, Figure 4B). Doubling the training volume used for delay and uncertainty estimation improved performance (rWIS 0.94, Figure 4B), while reducing it by half decreased performance (rWIS 1.08, Figure 4B). Applying the weekday filter method improved performance most of all the method specifications we explored (rWIS 0.93, Figure 4B).

**Figure 4:**
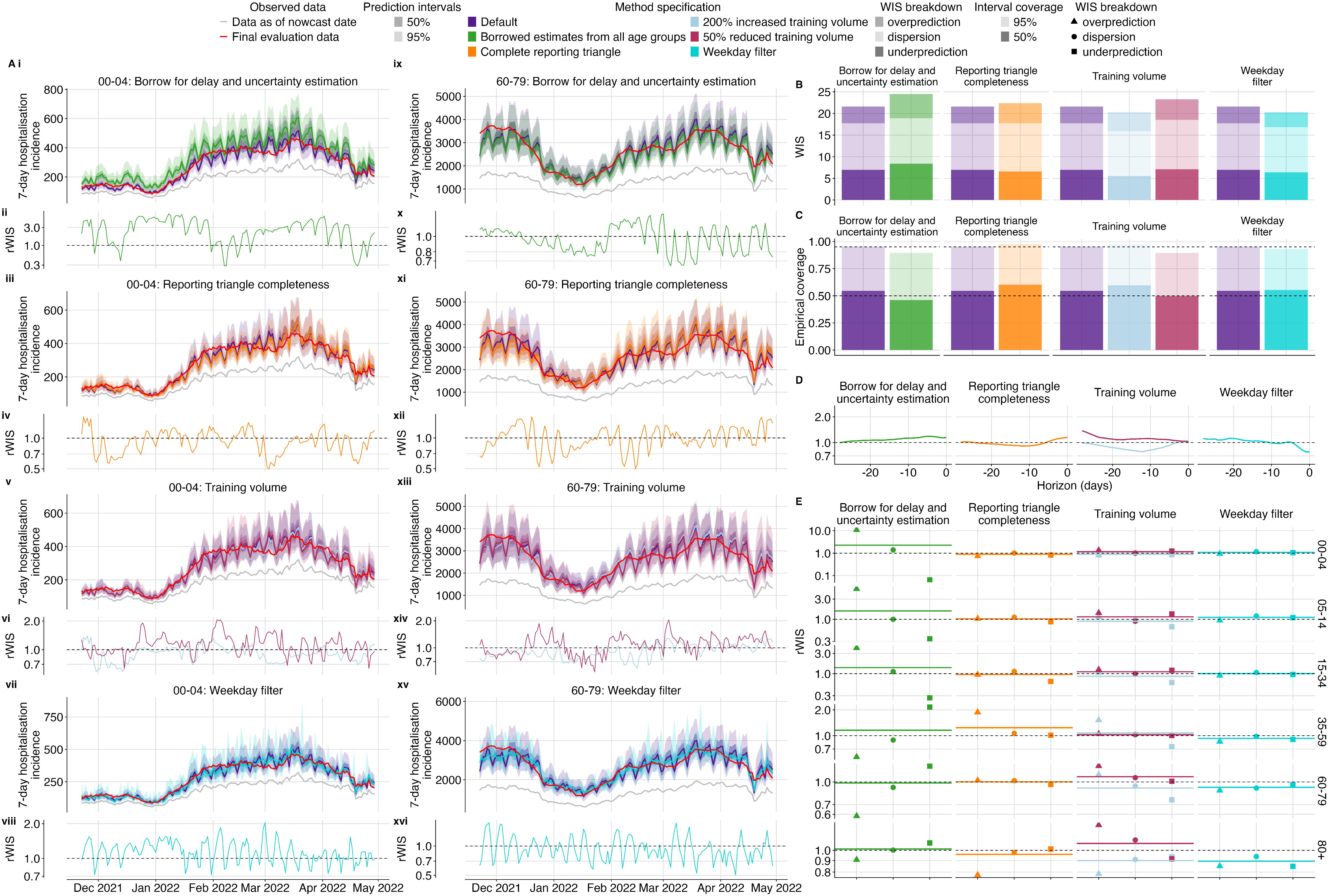
Performance comparison of different *baselinenowcast* method specifications compared to the default method. A. Upper plots show the daily same-day (0-horizon) nowcasts from the 00-04 (left) and 60-79 (right) age group for each method specification grouping showing the default validation model alongside variants from a specific permutation category: first row shows borrowing strategy variations (i, ix), second row shows reporting triangle completeness variations (iii, xi), third row shows training data volume variations (v, xiii), and the fourth row shows weekday filter variations (vii, xv). Observed data are shown as solid lines with gray indicating data available at the nowcast date and red indicating the final evaluation data. Colours indicate the method specification throughout, with the default approach in purple. Lower subplots show the relative WIS over time from the 00-04 (left) and 60-79 (right) age group relative to our default model with first row showing borrowing strategy variations (ii, x), second row showing reporting triangle completeness variations (iv, xii), third row showing training data volume variations (vi, xiv), and the fourth row showing weekday filter variations (viii, xvi). The horizontal reference line at 1.0 indicates performance parity with the validation default, values below 1.0 indicate improved performance of the method specification being considered. B. Overall performance comparison between all model configurations across all age groups, with decomposed absolute WIS (dispersion, overprediction, underprediction). Colour indicates method specification, with the default approach in purple. C. Empirical coverage for age group stratified nowcasts at 50% and 95% prediction intervals by method specification. Horizontal reference lines indicate the nominal coverage levels (50% and 95%). D. Relative WIS by nowcast horizon summarised across age groups. The horizontal reference line at 1.0 indicates performance parity with the default with values lower than 1.0 indicating improved performance of the method specification being considered. E. Relative decomposed WIS by age group, faceted by method specification grouping. Shape indicates relative WIS component (underprediction, dispersion, and overprediction).

Borrowing delay and uncertainty estimates from national data for each age group had the most pronounced reduction in performance in the sparsest age group (00-04) (rWIS 2.28), with the degradation in performance compared to the default specification driven by overprediction (Figure 4Ai, 4Aix, 4E, SI Figure 10). This reduction in performance when borrowing delay and uncertainty from all age groups is possibly due to the lower delay in the 00-04 age group compared to the overall delay across all age groups (Figure 3E, 3F). The greatest improvement amongst the age strata from borrowing delay and uncertainty estimates was modest, occurring in the 60-79 age group (rWIS 0.99)(Figure 4E, SI Figure 10). Borrowing strategies showed reduced coverage compared to the default (Figure 4C). Borrowing strategies also resulted in reduced performance at early horizons (Figure 4D) with the least available data, likely reflecting the reduced performance for the 00-04 age group at a horizon of 0 (Figure 4A). Averaging across age strata, performance was reduced due to borrowing for nearly all nowcast dates, except for at the beginning of February when we saw a slight improvement from borrowing (SI Figure 12), which might be due to the rapid fluctuation in the mean delay in the 05-14 age group a few weeks prior (Figure 4E).

Compared to the default, which uses the reporting triangle, visual inspection of nowcasts using only complete reporting matrices showed slightly improved performance in the 00-04 age group and degraded performance in the 60-79 age group (Figure 4Aiii, 4Aiv, 4Axi, 4Axii). The greatest improvement from using complete reporting matrices was in the 00-04 age group (rWIS 0.90, Figure 4Aiii, 4Aiv) and the greatest reduction in performance was in the 35-59 age group (rWIS 1.24) (Figure 4E, SI Figure 10). Across all age strata, complete reporting matrices demonstrated improved performance at the end of December 2021 and February 2022, with reduced performance elsewhere, suggesting improved performance of this specification during times of stable reporting delays (SI Figure 12). Reporting triangle completeness resulted in improvements near intermediate horizons -8 to -15 (Figure 4D) and increased coverage (Figure 4C).

Varying training data volume resulted in the most substantial differences in performance in the 00-04 age group which had the lowest counts. For this age group, doubling training volume showed improved performance (rWIS 0.89), while reducing the training volume by 50% reduced performance (rWIS 1.16)(Figure 4E, SI Figure 10). Visual inspection of the nowcast performance over time in the 00-04 and 60-79 year old age groups shows that trends in performance from increased and decreased training volume tend to oppose one another (Figure 4Av, 4Avi, 4Axiii, 4Axiv), indicating that there might not be a single optimal setting due to tradeoffs between overfitting to noise in reporting delays versus failing to incorporate the most recent temporal changes in reporting delay (SI Figure 12). Doubling the training volume resulted in improved performance at horizons between -8 and -15, whereas a 50% reduction in the training volume had considerably worse performance at distant horizons (-20 to -28) (Figure 4D). Increasing training data volume resulted in higher coverage (98% coverage), while reducing training volume reduced coverage (90% coverage), compared to the default (96% coverage) at 95% prediction intervals (Figure 4C).

Performing independent nowcasts of each weekday and combining them improved nowcast performance by reducing the large day-of-week variation observed in the default method (Figure 4Avii, 4Aviii, 4Axv, 4Axvi). Accounting for weekday effects in reporting had the greatest performance improvement in the most recent 7 horizon days (Figure 4D, SI Figure 9). Across age groups, the greatest performance improvement was observed in the 80+ age group (rWIS of 0.89), followed by the 60-79 age group (rWIS 0.92), with reductions in performance observed in the 00-04 and 05-14 age groups (rWIS of 1.09 and 1.11 respectively), which may be due to less of a consistent weekday effect in the sparser age groups compared to the larger age groups (Figure 3E).

### Case study: UKHSA norovirus surveillance

Visual inspection indicates that nowcasts from our base specification showed better alignment with eventual observed data compared to the naive baseline approach used in Mellor et al. [2] which was based on projecting the previous week’s data forward (Figure 5A, 5B, SI Figure 13). Comparing the external model implementations to our base specification, the epinowcast model had slightly better performance than our base specification (Figure 5E, 5F, SI Figure 13), with similarly high levels of uncertainty, while the GAM model showed reduced uncertainty and better overall performance compared to our base specification (Figure 5C, 5D, SI Figure 13).

**Figure 5:**
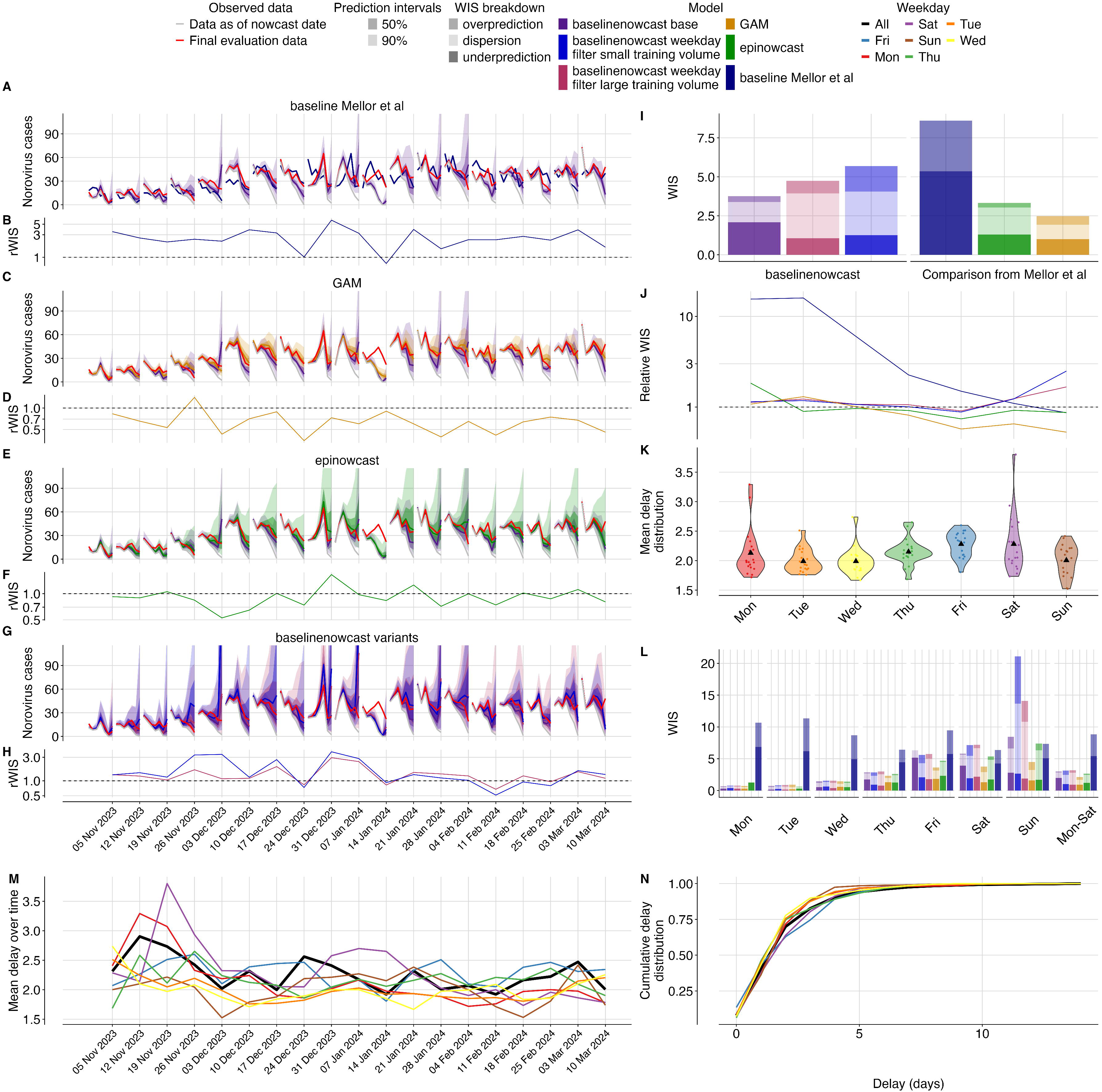
Nowcasting performance applied to UKHSA norovirus surveillance data. A-C-E-G. Weekly nowcasts compared to our base configuration, for three models used in Mellor et al. and compared to our two day-of-week filtered configurations. Observed data are shown as solid lines with gray indicating data available at the nowcast date and red indicating evaluation data. Nowcasts are shown out to 7 days as in Mellor et al. Shading indicates prediction intervals and colour indicates model, with the base specification in purple. B-D-F-H. Relative WIS over time (by week) compared to our base configuration. colours indicate models. Horizontal dashed line at 1.0 indicates performance parity with our base configuration, with value less than 1.0 indicating performance improvement of the method being considered. I. Overall performance comparison between all models, with decomposed absolute WIS (dispersion, overprediction, underprediction) displayed as stacked bar charts for direct comparison across our three variants and the three reference models from Mellor et al. Colour indicates model. J. Relative WIS by day of week for each model compared to our base specification with colour indicating model. Horizontal dashed line at 1.0 indicates performance parity with our base configuration. K. Distribution of mean reporting delays by weekday shown as violin plots with colours indicating the weekday. Black triangle indicates the mean delay for that weekday across nowcast dates. L. Mean reporting delays were calculated using the last 14 days of data (corresponding to the maximum delay). M. Mean reporting delay over time, with colours indicating the day of week and black indicating the mean reporting delay averaged across all weekdays. Mean reporting delays were calculated using the last 14 days of data (corresponding to the maximum delay). N. Cumulative distribution functions (CDFs) of the mean delay distribution. Colours indicate day of the week with black indicating the mean reporting delay averaged across all week days.

Overall, compared to our base specification, the Mellor et al. [2] baseline performed worse (rWIS 2.29), and both the the GAM model (rWIS 0.65) and the epinowcast implementation performed better (rWIS 0.89) (Figure 5I). The GAM model performed best relative to our base specification in December 2023 and late January to early February 2024 (Figure 5D), whereas the epinowcast implementation performed best relative to our base specification early December (Figure 5F). The absolute WIS of our base specification and the Mellor et al. models was consistent over time except for on January 14, 2024 (SI Figure 13), where all models performed poorly due to what appears to be initial underreporting in the most recent two days resulting in underpredictions by all models. Relatively speaking the GAM and epinowcast models both performed better than our base specification at this point (Figure 5D, 5F, SI Figure 13).

Our base specification and the GAM model were similarly calibrated (SI Figure 15) with slightly different coverage patterns. All models had 50% prediction intervals that were too wide with 69% (base specification), 59% (GAM), and 58% (epinowcast) coverage. At 90% prediction intervals, all models were well covered at 90% (base specification), 91% (GAM) and 88% (epinowcast) coverage (SI Figure 15). The Mellor et al. [2] baseline predictably undercovered for all prediction intervals and weekdays since it contained only a median estimate (SI Figure 15,16).

Performance varied substantially by day of week in the Mellor et al. models compared to our base specification (Figure 5J, 5L). As nowcasts were only conducted on Sundays, weekdays effectively reflect performance by nowcast horizon, though weekday effects in both specimen collection date and reporting also play a role. As expected, the most recent day with the least amount of data (Sunday, 0 horizon) showed the worst performance across models, whilst days further in the past showed progressively improved performance (Figure 5L). When comparing the GAM method and our base configuration, the most pronounced relative reduction in performance of our base specification was on Sunday, largely driven by reduced dispersion in the GAM compared to our base specification (Figure 5J, 5L), which had high dispersion on Sundays. Both the GAM and our method performed similarly for horizons -7 to -4 (weekdays Mon-Thurs, Figure 5J, 5L). Comparing the epinowcast implementation to our base specification revealed the largest relative improvement in performance of the epinowcast implementation was on Friday, when our base specification tended to underpredict (Figure 5L). Our base specification performed better than the epinowcast implementation for horizons -7 to -4 (weekdays Mon-Thur, Figure 5L).

Comparing our model variants to the base configuration, both weekday filter small and large training volume configurations reduced performance (rWIS 1.51 and 1.26 respectively) (Figure 5G, 5H, 5I). The visual differences were most notable in December 31, 2023 and January 7, 2024, where the weekday-filter variants appeared to exhibit more dispersion and overprediction than our base specification (Figure 5G, 5H, SI Figure 13). Overall, our base specification underpredicted whereas the weekday filter variations had higher dispersion and over prediction (Figure 5I). At 50% prediction intervals the weekday filter small and large training volume configurations had 74% and 76% coverage respectively compared to 58% coverage in our base specification (SI Figure 15). Our weekday-filter configurations performed less stably than our base model specification over time, with particularly poor relative performance of the weekday-filter with small training volume on November 26, 2023, and both configurations on December 31, 2023 and January 7, 2024 ( Figure 5G, 5H, SI Figure 13). Similar to the GAM and epinowcast models, the relative performance differences were most pronounced on Sunday. Both weekday-filter variants performed worse relatively on Sunday compared to our base specification (Figure 5J), but generally performed similarly on other days of the week (Figure 5J,L). The reduction in performance on Sunday appeared to be due in large part to dispersion for both weekday-filter variants, and in particular overprediction for the weekday-filter variant with a small training volume (Figure 5L).

## Discussion

Our baseline nowcasting method improved estimates of recent disease incidence compared to unadjusted data across all datasets evaluated. We verified that our *baselinenowcast* implementation was equivalent to nowcasts produced using a revised bug-fixed implementation of the original baseline method used in the German Nowcast Hub, and confirmed that it improved performance compared to the original implementation, highlighting the value of developing reproducible software for methods implementation to prevent the propagation of errors. As the original implementation of the KIT simple nowcast, despite the implementation bug, was competitive with more complex methods in German Nowcast Hub [3], ours is likely to have been even more competitive. Analysis of the performance of additional epidemiology-specific features such as weekday-specific delay estimation and strata sharing applied to the COVID-19 data showed how exploratory data analysis can help determine optimal baseline method specifications for the specific task. Our base specification substantially improved performance over the baseline used in the Mellor et al. [2] study, and helped to identify the horizons, weekdays, and nowcast dates in which the more complex models differed in performance compared to our baseline.

When we analysed the performance of different model specifications to understand tradeoffs and discover whether other specifications could improve nowcast performance in the context of the German COVID-19 data used in the Hub, we found modest improvements from applying a weekday filter to account for day of week effects in reporting and from increasing the amount of training volume used for delay and uncertainty estimation. Had these features been implemented in the German Nowcast Hub, this model would have been even more competitive, approaching the performance of the top performing model. This highlights the importance of exploratory data analysis to identify patterns, such as weekday effects and the time scale of changing reporting delays, to help determine optimal method specifications. We found reduced performance when deploying a strategy to borrow delay and uncertainty estimates from across all age groups. This approach reduced performance in the age groups with a greater difference in their average delay compared to the national average, as using a misspecified delay generally led to overprediction. However, at times of rapidly fluctuating reporting delays we would expect borrowing to be more beneficial as it will act to reduce noise in the delay estimate due to within-strata outliers.

In the UKHSA norovirus dataset, our model specifications fitting delay estimates independently to each weekday didn’t result in improved performance compared to our base configuration, likely because these methods required that we use a longer historical time period to have sufficient historical data for delay and uncertainty estimation. The reporting delay shifted enough over time to favor a more recent delay estimate, with less drastic weekday effects in delays than we had initially anticipated. These findings highlight that the distribution of delay estimates across strata, weekdays, and nowcast dates are key considerations when choosing model specifications, and can likely be used as a heuristic [39] to choose which model specifications are most appropriate in different epidemiological contexts.

Whilst we evaluated our approach across several datasets and through a range of method specifications, we did not consider a range of other possible method specifications. For example, users might also wish to use a different parametric distribution to model nowcast uncertainty. In this analysis we only considered negative binomial observation models applied to the target dataset, which in this case were always count data. Other valid error models can be readily explored, given the modular design of our baseline package. Additionally, users might wish to use our existing functionality in different combinations. For example, in this paper, we have only considered sharing information across all groups for both delay estimates and uncertainty, compared to independent estimates of both. A user might instead want to pool information for delay estimation but estimate uncertainty separately for each strata, or vice versa. We also only considered increasing training volume for both delay and uncertainty estimation together, and a user may wish to modify the combination of training data used for each task. A strength of our framework is that it supports these alternatives through its modular design.

For consistency and comparability with the original studies, we used the same evaluation metrics as those studies. This approach facilitated direct comparisons but meant we did not explore potentially more appropriate metrics, such as evaluating on log-transformed scales [40], which can more effectively capture relative differences when counts span multiple orders of magnitude, such as in the different age strata in the German Nowcast Hub COVID-19 data. Similarly, whilst WIS is a valid choice, we could have used Continuous Ranked Probability Score (CRPS), which scores probabilistic forecasts based on a full distribution of samples from the predictive distribution, avoiding information loss from using a limited set of quantiles. This was a particular issue in the Mellor et al. [2] comparison, where we followed their approach of using just two intervals (50th and 90th) when calculating WIS, where additional intervals may have better reflected the distribution of predictions. While this reduces the breadth of evaluation of the full predictive distribution, it has the benefit of focusing on evaluating only the prediction intervals that drive decision-making, namely those that are being visualised and presented to decision-makers. Another issue is that the use of retrospective datasets may not fully capture performance in truly prospective settings where evolving data availability and reporting changes create additional challenges. This is particularly true of the German COVID-19 data, where we used a final pre-processed reporting triangle for all analyses as vintages of that reporting triangle available each day were not systematically archived. However, we mitigated this issue by using case studies where data on reference and report times was available, as much as possible, as it would have been in real-time, and not tuning our models based on performance against these case studies. Additionally, our focus on proper scoring rules provides a standardised assessment but might not capture all dimensions of nowcast quality relevant to decision-making [41]. Finally, the datasets selected, while diverse, do not represent the full spectrum of delay structures and reporting processes encountered in public health surveillance.

Multiple methods have been proposed as nowcasting approaches in infectious disease surveillance. Most of these methods jointly estimate reporting delays and the final number of cases [18,21,22,26,42], though other methods for nowcasting are similar to ours in that they require specifying an independently estimated delay distribution [43]. The joint approach has the advantage that the full set of training data is used to estimate both delays and uncertainty, whereas our baseline method requires use of a proportion of the training data for delay estimation, so that we have sufficient data to estimate uncertainty using the same amount of training data applied at different retrospective nowcast times. Other methods differ in how they model the time evolution of the final number of cases, the delay distribution, and the observation model. For example, some assume a parametric form for the reporting delay distribution, while others assume a non-parametric delay. A few methods enable estimation of time-varying delays [7,42]. Commonly, hazard-based formulations are used [18,21,44]. Methods also take varying approaches to smoothing, including the use of geometric random walks [21,26,42] and B-splines [22]. Individual methods have also been extended to support modelling day of the week variation, covariate inclusion, public holiday anomalous reporting, and transmission processes [4,6,7,19,20]. Bayesian methods [6,7,18,19,26,44,45] can be particularly useful in settings with sparse data, where the ability to specify priors and expected reporting patterns is critical, for example, in novel outbreaks or situations with very low case counts. However, many of these methods are relatively complex which can make them difficult to inspect, validate, or extend, and may make it unclear when these models are inappropriate and prone to failure due to reasons such as unidentifiability or non-convergence. While other domains have implementations of a similar multiplicative approach to ours, such as the *chainladder* R [16,17] package designed for nowcasting insurance claims, we have not seen widespread adoption likely due to the lack of examples in epidemiological contexts or the addition of features specific to the infectious disease context.

In contrast, our approach focuses on a minimal set of well-defined, modular components that are simple and easy to explain. The method has a modular, pipe-friendly design enabling users to customise specific components. By breaking the nowcasting method into distinct components, users can adapt, inspect and validate each part of the model. This simplicity makes implementation and model fitting straightforward without requiring specialist knowledge, making it suitable for implementation in public health agencies with moderate statistical expertise. A key advantage of our method over similar methods, such as the chainladder R package [17], is simply that it is tailored to common epidemiological contexts; with its modular design enabling the generation of nowcasts for different temporal granularities in reference and report dates, ease and demonstration of how to “borrow” delays and uncertainty across different strata, and the ability to independently fit each weekday to account for weekday effects in reporting, all of which are common scenarios encountered in epidemiology and which *baselinenowcast* was designed to handle. Additionally, based on retrospective performance, the uncertainty quantification approach provides relatively well-calibrated nowcasts. We note this was not the case for many of the models submitted to the original German Nowcast Hub [3] and we consider this to be a key advantage of this baseline approach over other baseline methods such as the method used by Mellor et al. [2] which propagated the previous week’s data forward with no uncertainty. Lastly, by providing a standardised R package with comprehensive documentation, our method aims to address challenges in reproducing and re-implementing different baseline methods, ensuring that analyses applied in one context can be readily extended to other pathogens/surveillance systems with relative ease and standardized approaches.

However, important limitations in our method exist. The method follows a pipeline approach where delay distributions are estimated independently from case occurrence processes. As demonstrated by [5], such approaches can propagate errors and may not fully account for interdependencies between these processes. The method also lacks flexibility for incorporating time-varying delay distributions or partially pooled estimates across strata, which may be crucial during periods of changing surveillance intensity or for rare diseases where flexibly borrowing information is essential.

Our approach to handling day-of-week effects highlights an important trade-off. By filtering data to use only observations from the same day of the week, we provide a simple solution that avoids complex modelling whilst effectively capturing weekly patterns. However, this simplicity comes at the cost of discarding potentially valuable data on recent trends in delay estimation, which proved to be problematic when performing independent estimates of the delay across a large time frame in the norovirus case study. More sophisticated approaches, such as those incorporating day-of-week terms in statistical models [19], as was used by the GAM implementation in [2], can utilise all available data but require greater statistical complexity. This illustrates a broader tension in nowcasting between accessible methods that may sacrifice efficiency and complex methods that maximise statistical power but may be less accessible to practitioners.

More fully featured nowcasting frameworks, such as the *epinowcast* R package (a semi-mechanistic Bayesian approach [19], which was used in the Mellor et al study [2]), and generalised additive model-based approaches [4], encompass the functionality of other nowcasting approaches whilst offering greater flexibility, often via formula interfaces, and standardised tooling. This increased flexibility allows users to customise models for their specific problem reflecting their familiarity with details of the data and how they are collected, but has the downside of increasing model, software, and interface complexity. The evidence supporting the benefits of this increased flexibility is currently limited, as little work has been done to evaluate framework-based approaches.

Future research should focus on five key areas. First, developing standardised benchmarks and datasets would enable rigorous evaluation of nowcasting methods, quantifying trade-offs between simplicity and performance across different surveillance contexts. Second, research addressing practitioners’ needs through decision support tools could guide method selection based on data characteristics and available resources. Relatedly, co-creation of documentation with public health practitioners, as well as evaluation in the real-world public health context will be critical to help address implementation barriers. Thirdly, research is needed to implement a wider array of error models for improved uncertainty quantification. Fourthly, reporting processes can be complex, for example they can include upwards revisions or spatial drop out in reporting. Both the type of empirically based methods we discuss in this work and more complex methods need to be adapted for these settings. Finally, advancements in model composition could enable seamless transitions between nowcasting and forecasting and address error propagation issues inherent in pipeline methods [5,46].

In conclusion, our baseline nowcasting method addresses gaps in the epidemiological toolbox by providing both a practical nowcasting solution for infectious disease surveillance and a standard benchmark for further model development. Its performance across datasets demonstrates that straightforward approaches can provide substantial improvements over unadjusted data whilst remaining accessible to practitioners. Our discovery of the implementation issue in the KIT simple nowcast method highlights the value of implementing reproducible, tested, and validated software. The performance of different method specifications highlighted the importance of exploratory data analysis to identify patterns in the delay distributions across time and strata. The method’s pipe-friendly, modular design enables simple yet effective adaptations, such as the day-of-week stratification, without requiring complex statistical frameworks. Our method, developed with public health practitioners, is robust, interpretable, and relatively straightforward. By establishing this benchmark, we aim to reduce the barriers to implementing nowcasting solutions in public health practice and provide a resource for improving nowcasting methods, ultimately contributing to more timely and effective public health responses.

## Data & software availability statement

Code to reproduce results and figures are provided at https://github.com/epinowcast/baselinenowast-paper with a stable release published at 10.5281/zenodo.16792463. Functions used to generate nowcasts are available as in the baselinenowcast package at https://github.com/epinowcast/baselinenowcast with a stable release published at 10.5281/zenodo.16792456. Input data used to generate COVID-19 nowcasts from Germany is available at https://github.com/KITmetricslab/hospitalization-nowcast-hub. Data on norovirus cases in England anonymized and with statistical noise added is available at https://github.com/jonathonmellor/norovirus-nowcast-baselinenowcast.

## AI disclosure

Generative AI tools were used to assist with preparation of the manuscript text and as part of code review for both the *baselinenowcast* R package and the analysis applying it to the two case studies presented here. Coderabbitai https://github.com/apps/coderabbitai was used for automated code review. Claude Pro version 4 https://claude.ai/ was used for assistance with writing code and for revisions of text in the manuscript. Claude Sonnet 4 https://www.anthropic.com/claude/sonnet was used for writing and revising code. Opus https://opus.ai/ was used for assistance with writing.

## Author contributions

**Conceptualization:** Kaitlyn E Johnson, Sam Abbott, Sebastian Funk.

**Data curation:** Johannes Bracher, Daniel Wolffram, Jon Mellor, Maria L Tang.

**Formal analysis:** Kaitlyn E Johnson, Sam Abbott, Maria L Tang, Jon Mellor.

**Funding Acquisition**: Nicholas Reich, Sebastian Funk.

**Investigation:** Kaitlyn E Johnson, Sam Abbott.

**Methodology:** Johannes Bracher, Kaitlyn E Johnson, Sam Abbott, Sebastian Funk.

**Project administration:** Sam Abbott, Sebastian Funk, Kaitlyn E Johnson.

**Resources:** Sebastian Funk

**Software:** Kaitlyn E Johnson, Sam Abbott.

**Supervision:** Sam Abbott, Sebastian Funk.

**Validation:** Kaitlyn E Johnson, Sam Abbott, Maria L Tang, Jon Mellor.

**Visualization:** Kaitlyn E Johnson, Sam Abbott, Emily Tyszka, Laura Jones, Rosa Ergas

**Writing-original draft:** Kaitlyn E Johnson, Sam Abbott.

**Writing-reviewing & editing:** Kaitlyn E Johnson, Sam Abbott, Sebastian Funk, Johannes Bracher, Laura Jones, Emily Tyszka, Maria L Tang, Jon Mellor, Rosa Ergas, Nicholas Reich, Barbora Nemcova.

