## Supplement for "Baseline nowcasting methods for handling delays in epidemiological data"

#### Supplemental methods

##### Mathematical model

Our approach is based on the reference model for the COVID-19 hospitalisation nowcast challenge in Germany in 2021 and 2022 (1). In slight variation of the chain ladder method (2), it uses preliminary case counts and empirical delay distributions to estimate yet-to-be-observed cases. Probabilistic nowcasts are generated using a negative binomial model with means from the point nowcast and dispersion parameters estimated from past nowcast errors. Below, we describe the mathematical details of each component of the model, starting with a definition of the notation used throughout. In our description we assume that time steps correspond to days, but they could also be weeks, months or any other unit.

See the *Default Settings* section in the main text methods for a description of the default behaviour of the method within the `baselinenowcast` package.

##### Notation

We denote by $X_{t,d}, d = 0,..,D$ the number of cases occurring on day $t$ which appear in the dataset with a delay of $d$ days. For example, a delay $d = 0$ means that a case occurring on day $t$ arrived in the dataset on day $t$. We only consider cases reported within a maximum delay $D$. The number of cases reporting for time $t$ with a delay of at most $d$ can be written as:

$$X_{t,\leq d} = \sum_{i=0}^{d} X_{t,i} \qquad (1)$$

A special case of this is the "final" number of reported cases at time $t$, denoted by

$$X_t = X_{t,\leq D} = \sum_{i=0}^{D} X_{t,i}.$$

For delays $d < D$ we moreover define the notation

$$X_{t,>d} = \sum_{i=d+1}^{D} X_{t,i}$$

representing the number of cases still missing after $d$ days of delay. In the following we use uppercase letters ($X_t$) for random variables, lower case ($x_t$) for the corresponding observations, and hats ($\hat{x}_t$) for estimated / imputed values. The matrix $\mathbf{x}$ with entries $x_{t,d}, t = 1, \dots, t^*, d = 1, \dots, D$ is referred to as the *reporting matrix*:

| | $d = 0$ | $d = 1$ | $d = 2$ | … | $d = D - 1$ | $d = D$ |
|---|---|---|---|---|---|---|
| $t = 1$ | $x_{1,0}$ | $x_{1,1}$ | $x_{1,2}$ | … | $x_{1,D-1}$ | $x_{1,D}$ |
| $t = 2$ | $x_{2,0}$ | $x_{2,1}$ | $x_{2,2}$ | … | $x_{2,D-1}$ | $x_{2,D}$ |
| $t = 3$ | $x_{3,0}$ | $x_{3,1}$ | $x_{3,2}$ | … | $x_{3,D-1}$ | $x_{3,D}$ |
| … | … | … | … | … | … | … |
| $t = t^* - 2$ | $x_{t^*-2,0}$ | $x_{t^*-2,1}$ | $x_{t^*-2,2}$ | … | $x_{t^*-2,D-1}$ | $x_{t^*-2,D}$ |
| $t = t^* - 1$ | $x_{t^*-1,0}$ | $x_{t^*-1,1}$ | $x_{t^*-1,2}$ | … | $x_{t^*-1,D-1}$ | $x_{t^*-1,D}$ |
| $t = t^*$ | $x_{t^*,0}$ | $x_{t^*,1}$ | $x_{t^*,2}$ | … | $x_{t^*,D-1}$ | $x_{t^*,D}$ |

In the case where $t^*$ corresponds to the present date, all entries with $t + d > t^*$ have yet to be observed and are thus still missing. As the available entries at its bottom form a triangle, this incomplete reporting matrix is referred to as the *reporting triangle*.

| | | | | |
|---|---|---|---|---|
| $t = t^* - 2$ | $x_{t^*-2,0}$ | $x_{t^*-2,1}$ | $x_{t^*-2,2}$ | … |
| $t = t^* - 1$ | $x_{t^*-1,0}$ | $x_{t^*-1,1}$ | | … |
| $t = t^*$ | $x_{t^*,0}$ | | | … |

#### Pre-processing of the reporting triangle

All of the following steps require that the reporting triangle only has non-negative entries. In practice this is not necessarily the case. For instance, if the reporting triangle has been computed from increments in subsequent data snapshots, occasional downward corrections due to data entrance issues can cause negative entries. We therefore apply a pre-processing step to re-distribute negative entries across neighbouring cells with positive entries.

#### Delay distribution estimation

In this section we detail how we estimate the reporting delay distribution based on the last $N$ rows of a reporting matrix or triangle. See the Methods: Overview section in the main text for more details on the package default for the number of reference times used for delay estimation.

*Estimating the delay distribution from a reporting matrix*

If a complete reporting matrix is available, estimating the discrete-time delay distribution $\pi_d, d = 0, \ldots, D$ is straightforward. Using the last $N$ rows of the reporting matrix $\mathbf{x}$, we compute

$$\hat{\pi}_d = \frac{\sum_{t=t^*-N+1}^{t=t^*} x_{t,d}}{\sum_{t=t^*-N+1}^{t=t^*} x_t}, \qquad (2)$$

which is simply the relative frequency of a delay of $d$ days among all cases in the reporting matrix.

*Estimating the delay from a reporting triangle*

In the case where $t^*$ is the present day, such that only a reporting triangle with missing entries is available, the estimator $\hat{\pi}_d$ from Equation 2 can only be evaluated after discarding all data from the last $D - 1$ time points. In order to use these partial observations, we use a different representation of the delay distribution via terms of the form

$$\theta_d = \frac{\pi_d}{\pi_{\leq d-1}} \qquad (3)$$

for $d = 1, \ldots, D$. Here, in analogy to Equation 1 we write

$$\pi_{\leq d-1} = \sum_{d\prime=0}^{d-1} \pi_d\,. \qquad (4)$$

The $\theta_d$ can be estimated via

$$\hat{\theta}_d = \frac{\sum_{t=t^*-N+1}^{t^*-d} x_{t,d}}{\sum_{t=t^*-N+1}^{t^*-d} x_{t,\leq d-1}},$$

and translated to estimates $\hat{\pi}_0, \ldots, \hat{\pi}_D$ via the recursion

$$\hat{\pi}_{\leq d} = \left(1 + \hat{\theta}_d\right)\hat{\pi}_{\leq d-1}$$

subject to the constrain that $\sum_{d=0}^{D} \pi_d = 1$.

We note that this method is equivalent to the so-called chainladder method (2), adapted to our notation in terms of reporting triangles (rather than *development triangles* as used in accounting).

#### Point nowcast generation

We now address the computation of a point nowcast, i.e., expected total daily case numbers $\hat{x}_t, t = t^* - D + 1, t^*$. These are based on the reporting triangle, more specifically the preliminary totals $x_{t,\leq t^*-t}$, and the estimated delay distribution, $\hat{\pi}_d, d = 0, \ldots, D$. In the following we will denote the current day $t^*$ as the nowcast date, and the days $t = t^* - D, \dots, t^*$ as the reference dates. The difference $j = t^* - t$ will be called the *horizon*.

An intuitive approach, used in (1) and the standard chain ladder technique, is to simply inflate the current total for a reference date $t$ by the inverse of the respective probability of observation up to time $t^*$,

$$\hat{x}_t = \frac{x_{t,\le t^*-t}}{\pi_{\le t^*-t}}.$$

This, however, is not well-behaved if no cases at $t$ have been observed yet, i.e., $x_{t,\le t^*-t} = 0$. Then $\hat{x}_t$ is likewise zero, which yields problems in our uncertainty quantification method (see next section). Motivated by a Bayesian argument (see Zero-handling below) we therefore use the expression

$$\hat{x}_t = \frac{x_{t,\le t^*-t} + 1 - \pi_{\le t^*-t}}{\hat{\pi}_{t,\le t^*-t}} \qquad (5)$$

instead. This yields essentially identical results for large $x_{t,\le t^*-t}$, but produces positive $\hat{x}_t$ even for preliminary zero values $x_{t,\le t^*-t} = 0$.

For our uncertainty quantification scheme we require not only estimated totals $\hat{x}_t$, but all entries $\hat{x}_{t,d}$ of a point nowcast matrix. For $t = t^* - D + 1, \dots, t^*, d > t^* - t$ these are obtained as

$$\hat{x}_{t,d} = \hat{\pi}_d \times \hat{x}_t.$$

#### Uncertainty quantification

To estimate the uncertainty in the nowcasts, we use the nowcast errors from $M$ past nowcasting time points. See the Methods: Overview section for more details on the default settings used in the package to define the number of $M$ past nowcasting time points used.

##### *Generation of retrospective reporting triangles*

We first obtain "vintage" reporting triangles of the raw reporting triangle (i.e., before pre-processing) to replicate the data which would have been available as of times $s^* = t^* - 1, \dots, t^* - M$, i.e., the last $M$ days on which nowcasts could have been generated. This simply corresponds to the stepwise omission of all entries with $t + d > s^*$, which for each $s^*$ is a diagonal from the bottom left to the top right. The same pre-processing step as in Section Preprocessing of the reporting triangle is applied to each vintage reporting triangle.

##### *Generation of retrospective point nowcast matrices*

For each of the $M$ vintage reporting triangles, i.e., $s^* = t^* - 1, , \dots, t^* - M$, we apply the method described above to estimate a delay distribution and generate a point nowcast matrix. To indicate the data version on which it is based, its entries are denoted by

$$\hat{x}_{t,d}(s^*)$$

for $t = s^* - D + 1, \ldots, s^*$ and $d = s^* - t + 1, \ldots D$.

Note that estimation is again based on the last $N$ rows of the respective reporting triangle, which must consequently contain at least $M + N$ rows in total.

##### *Fit a negative binomial observation model to past nowcast errors (per nowcast horizon)*

A point nowcast based on the reporting triangle from time $s^*$ can also be written as

$$\hat{x}_t(s^*) = x_{t,\leq s^*-t} + \hat{x}_{t,>s^*-t}(s^*).$$

Only the second term has some associated uncertainty, while the first is already known at time $s^*$. To quantify this uncertainty for given nowcast horizon $0 \leq j \leq D$, we assemble the $\hat{x}_{s^*-j,>j}(s^*)$ and $x_{s^*-j,>j}$ for $s^* = t^* - M, \ldots, t^* - 1$. If all observations were complete, we would then estimate the overdispersion parameter $\phi_j$ of a negative binomial distribution

$$X_{s^*-j,>j} \sim \text{NegBin}\left[\hat{x}_{s^*-j,>j}(s^*), \phi_j\right], \qquad (6)$$

with independence assumed across the different $s^*$. This, however, is not directly feasible as again some of the $x_{s^*-j,>j}$ are not yet observed at time $t^*$. We could discard these instances, but this would considerably reduce our number of available observations. We therefore use partial observations as available at time $t^*$ and assume

$$\left(\sum_{d=j+1}^{\min(D,t^*-s^*)} X_{s^*-j,d}\right) \sim \text{NegBin}\left[\sum_{d=j+1}^{\min(D,t^*-s^*)} \hat{x}_{s^*-j,d}\,, \phi_j\right]. \qquad (7)$$

Here, the use of a constant dispersion parameter $\phi_j$ despite some of the values in the fitting procedure being yet incomplete is justified by the fact that the negative binomial distribution is closed to binomial subsampling, with the overdispersion parameter preserved. If equation Equation 6 holds in combination with

$$\left(\sum_{d=j+1}^{\min(D,t^*-s^*)} X_{s^*-j,d}\right) \mid X_{s^*-j,>j} \sim \text{Bin}\left[X_{s^*-j,>j}, \left(\sum_{d=j+1}^{\min(D,t^*-s^*)} \pi_d\right)/\pi_{>j}\right],$$

we thus obtain Equation 7. Estimated dispersion parameters $\hat{\phi}_0, \ldots, \hat{\phi}_D$ are obtained by maximum likelihood estimation.

#### Probabilistic nowcast generation

Predictive distributions for $X_{t^*}, \ldots, X_{t^*-D+1}$ are obtained by combining our point nowcasts with the estimated overdispersion parameters in a simple plug-in fashion. Specifically, we set

$$X_{t,>t^*-t} \sim \text{NegBin}\left(\hat{x}_{t,>t^*-t}, \phi_{t^*-t}\right).$$

The predictive distribution for $X_t$ then results by shifting this distribution by the already known value $x_{t,\leq t^*-t}$.

#### Zero-handling strategy

As mentioned in Point nowcast generation, we use a modified point nowcasts to deal with zero values in preliminary counts. We here motivate this approach from a Bayesian perspective, based on the work of (3). To this end we assume that

$$X_{t,\leq d} \mid X_t \sim \text{Bin}\left(X_t, \sum_{d=0}^{d} \pi_t\right).$$

We are now interested in the conditional expectation

$$\mathbb{E}(X_t \mid X_{t,\leq d})$$

in this binomial subsampling problem. We will derive it under the improper prior distribution

$$X_t \sim \text{DiscreteUniform}(0,1,2,\ldots).$$

For notational simplicity and readability for the following, we substitute $N = X_t$, $Y = X_{t,\geq d}$ and $p = \sum_{d=0}^{D} \pi_d$ and are thus looking for $\mathbb{E}(N|Y = y)$ if $Y \sim \text{Bin}(N, p)$ with a discrete uniform prior on $N$. This expectation can be written out as:

$$\mathbb{E}(N \mid Y = y) = \sum_{n=0}^{\infty} \Pr(N = n|Y = y) \times n \qquad (8)$$

Applying Bayes Theorem we have

$$\Pr(N = n \mid Y = y) = \frac{\Pr(Y = y \mid N = n) \times \Pr(N = n)}{\sum_{i=0}^{\infty} \Pr(Y = y \mid N = i) \times \Pr(N = i)}.$$

Because $\Pr(N = n)$ is a constant this simplifies to

$$\Pr(N = n \mid Y = y) = \frac{\Pr(Y = y \mid N = n)}{\sum_{i=0}^{\infty} \Pr(Y = y \mid N = i)}.$$

Now substituting the probability mass function of the binomial distribution

$$\Pr(Y = y \mid N = n) = \binom{n}{y} p^y (1-p)^{n-y}$$

we get

$$\Pr(N = n \mid Y = y) = \frac{\binom{n}{y} p^y (1-p)^{n-y}}{\sum_{i=1}^{\infty} \binom{1}{y} p^y (1-p)^{i-y}}$$

Plugging this into  we get the following (omitting terms for $n < y$, which are 0):

$$\mathbb{E}(N \mid Y = y) = \sum_{n=y}^{\infty} n \frac{\binom{n}{y} p^y (1-p)^{n-y}}{\sum_{i=y}^{\infty} \binom{i}{y} p^y (1-p)^{i-y}}$$

This is equivalent to

$$\mathbb{E}(N \mid Y = y) = \frac{\sum_{n=y}^{\infty} n \binom{n}{y} p^y (1-p)^{n-y}}{\sum_{i=y}^{\infty} \binom{i}{y} p^y (1-p)^{i-y}}.$$

Both the numerator and the denominator are known convergent series, with solutions available in standard libraries like Mathematica (4). We then get

$$\mathbb{E}(N \mid Y = y) = \frac{(y+1-p)/p^2}{1/p} = \frac{y+1-p}{p},$$

which corresponds to the corrected point estimate provided in equation Equation 5.

#### Description of KIT simple nowcast implementation issue and revised nowcasts

When verifying the KIT simple nowcast implementation, we noticed that the code used to generate the KIT simple nowcasts in real-time contained an additional indexed element in the delay PMF for all counts observed beyond the maximum delay thus far. In the generation of retrospective nowcasts, these values were being handled as having been observed as 0s, when in fact they had yet to be observed in all of the retrospective nowcasts. This resulted in a comparison of a 0 to a value that in some cases is eventually observed, rather than beign excluded as they should have been. This has the impact of inflating the dispersion estimates, and may explain why the KIT simple nowcast method in Wolffram et al. (1) overcovers relative to the other methods. In the latest implementation in the RESPINOW Hub (5), the authors have removed the density beyond the maximum delay column from their implementation, and thus it no longer contains this issue.

In order to validate our `baselinenowcast` method against a revised version with the bug fixed, we regenerated the nowcasts using the pre-processed reporting triangle in the German Nowcast Hub repository (6) and generated retrospective nowcasts, referring to these in the main text and supplement as the KIT simple nowcast revised. To facilitate a fair comparison to the original implementation, we also reran the original implementation on the same pre-processed reporting triangle and present those results below, with the original implementation labeled as “KIT simple nowcast original:.

### Additional figures

#### German COVID-19 Nowcast Hub validation: using the original KIT simple nowcast implementation

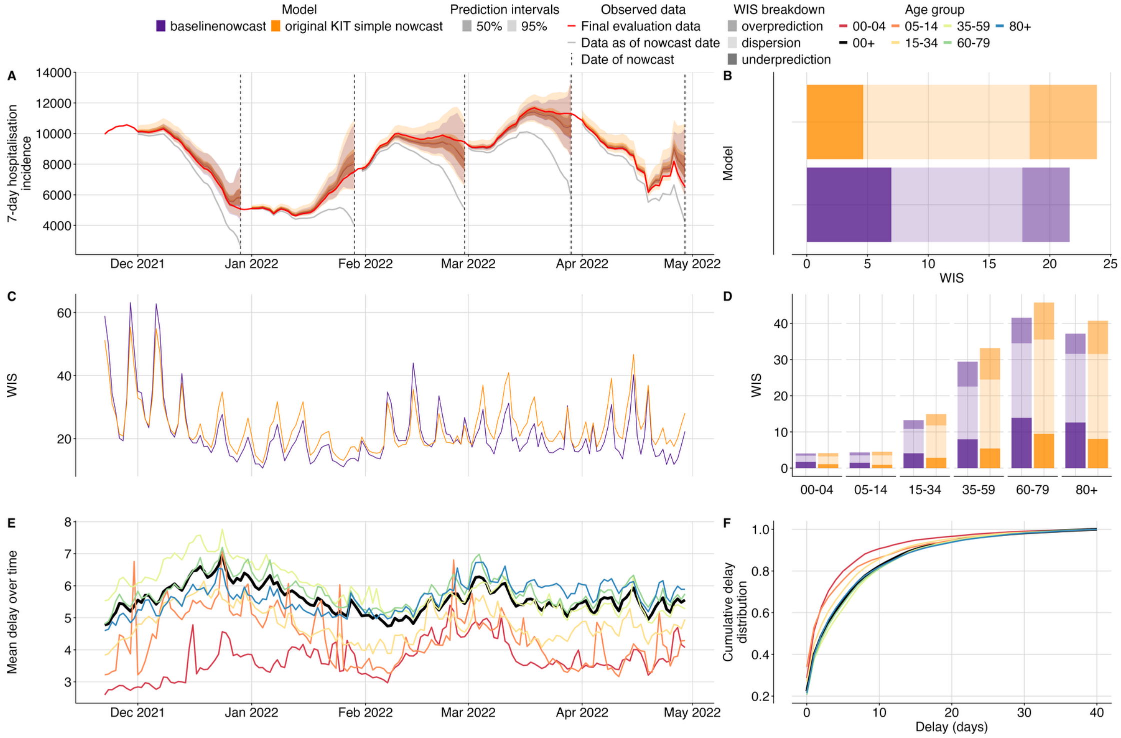


*Fig. S1 Validation of baseline nowcasting model using German COVID-19 data compared to original implementation of the KIT simple nowcast. A. Illustration of the nowcasting task from 5 nowcast dates throughout the study period. Nowcasts from our baseline model (purple) and the original implementation of the KIT simple nowcast model plotted against eventually observed values aggregated across all age groups, coloured by model. Observed data are shown as solid lines with gray indicating data available at the nowcast date and red indicating the final evaluation data. Dashed vertical lines indicate the date of the nowcasts. Shading indicates prediction intervals of each nowcast. Nowcasts are shown from horizons 0 to -28 days. B. Overall performance comparison between models aggregated across performance in individual age strata, with decomposed WIS (dispersion, overprediction, underprediction) displayed as stacked bar charts with different shading for direct comparison. C. Performance over time, displaying daily mean WIS scores summarised across age strata and 28 horizon days, coloured by model. D. Performance by age group, showing WIS scores across different age groups on the x-axis with grouped bars for both models and stacked shaded components displaying the decomposition of scores. E. Mean reporting delay over time, visualised as multiple coloured lines representing each age group, with the national average shown as a thicker black line. Mean reporting delays were calculated using the last 40 reference dates, corresponding to the maximum delay, for each nowcast date and age group. F: Delay distribution presented as cumulative*

*distribution functions (CDFs), with overlaid curves for each age group and the national average highlighted as a thicker black line.*

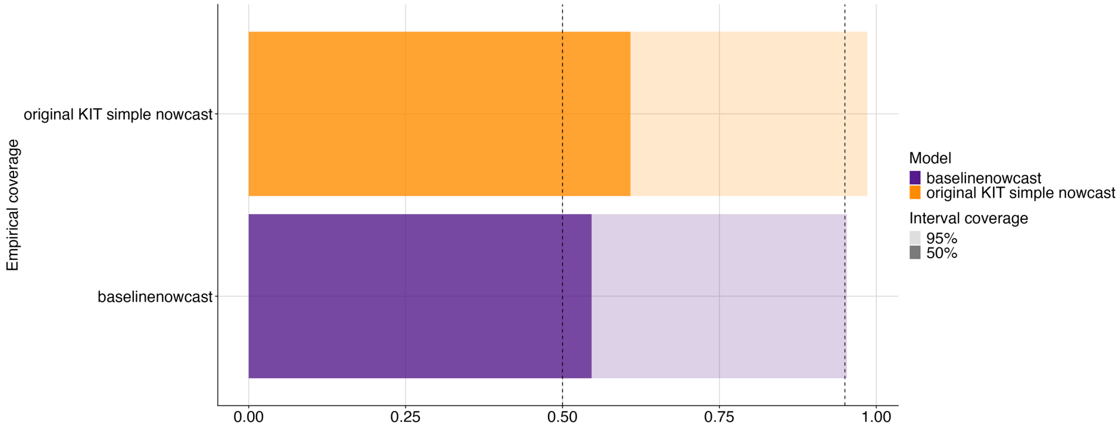


*Fig. S2 Empirical coverage of baselinenowcast method and real-time KIT simple nowcast at 50% and 95% prediction intervals. Shading indicates prediction intervals, colours indicate model.*

#### German Nowcast Hub validation

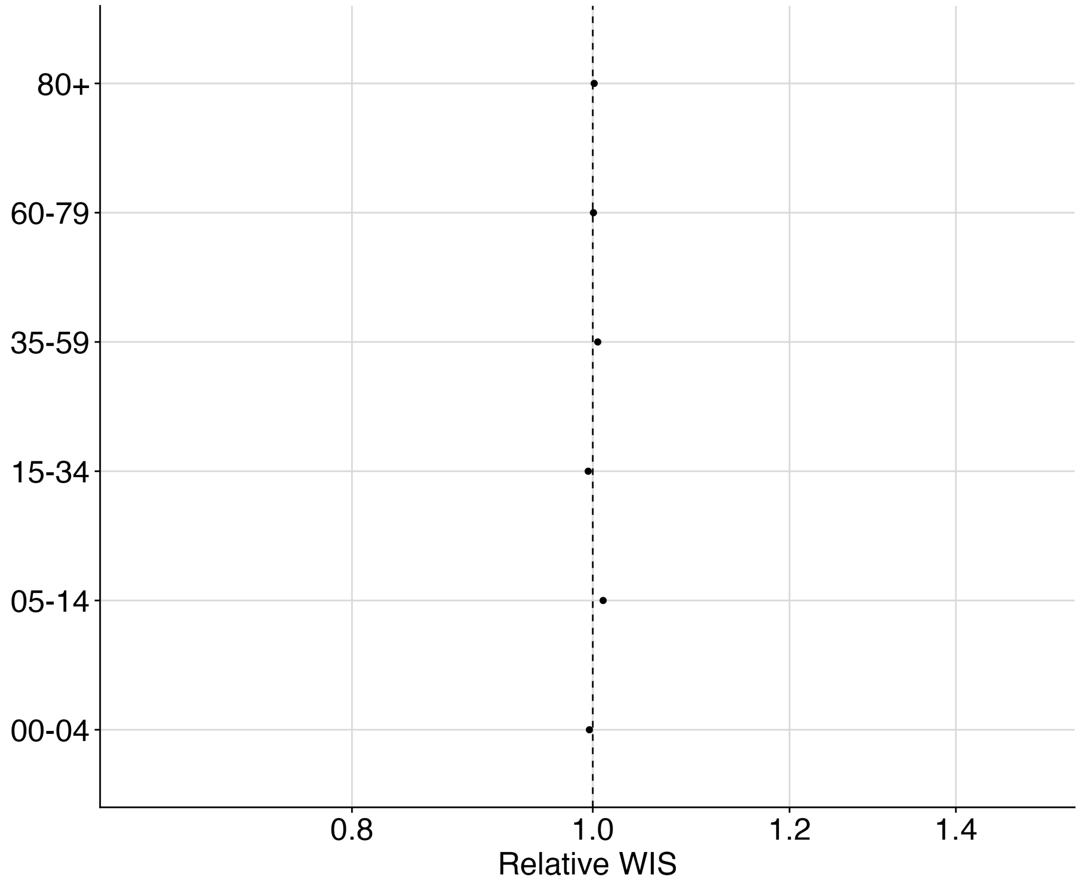


*Fig. S3 Relative WIS by age group of baselinenowcast compared to KIT simple nowcast revised. Vertical line at 1.0 indicates parity with KIT simple nowcast revised.*

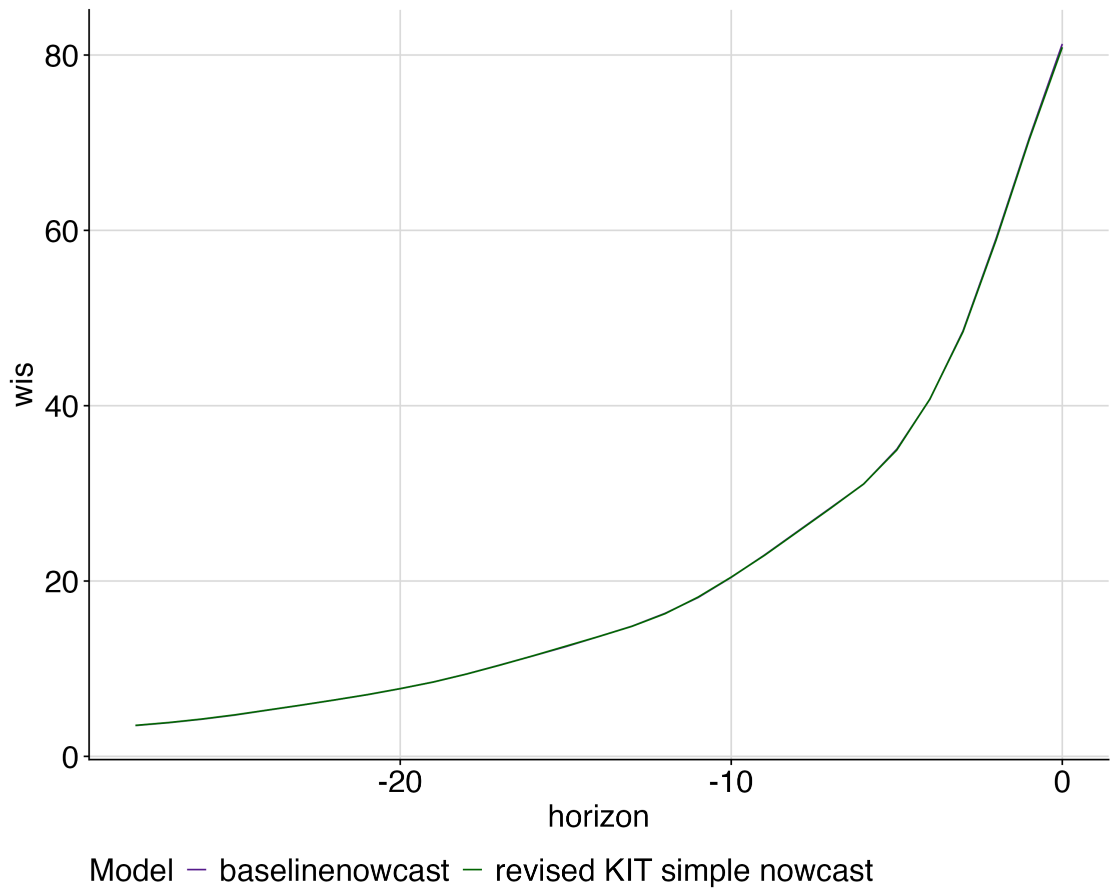


*Fig. S4 Mean WIS by nowcast horizon for each model. colours indicate models.*

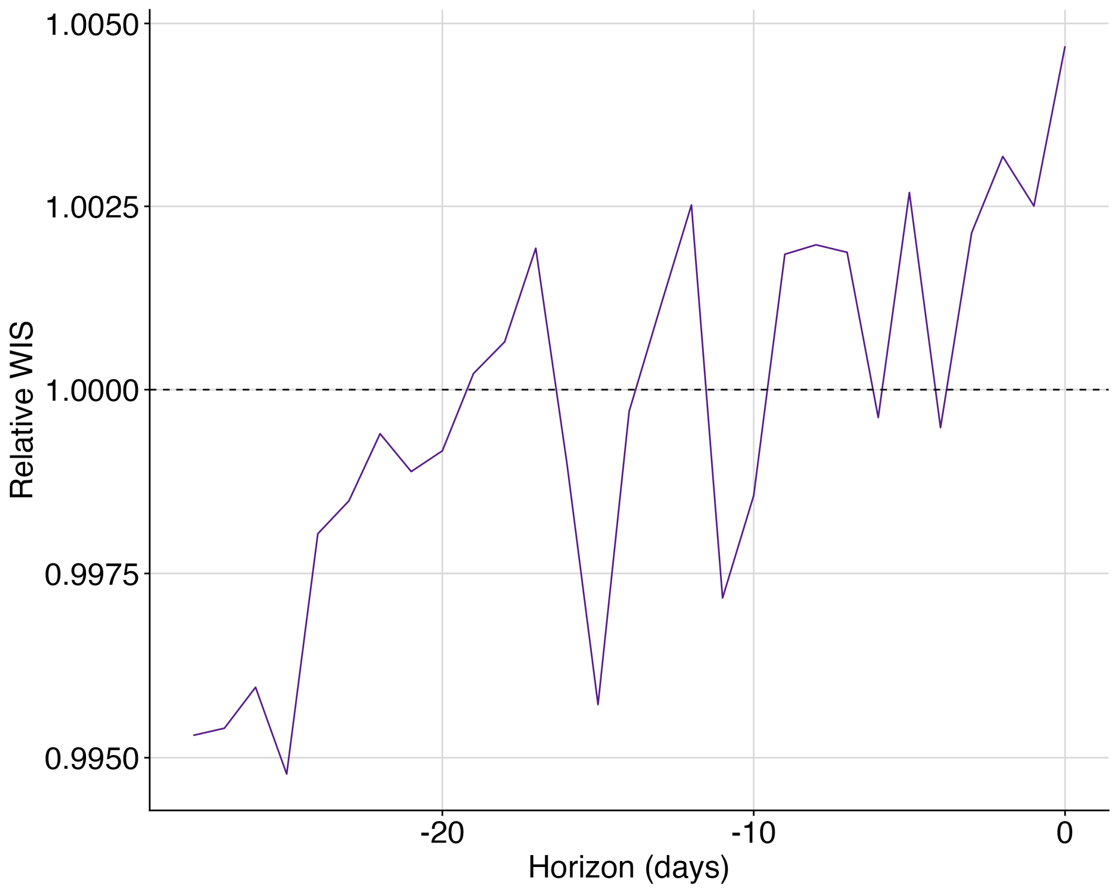


*Fig. S5 Relative WIS of baselinenowcast horizon compared to KIT simple nowcast revised.*

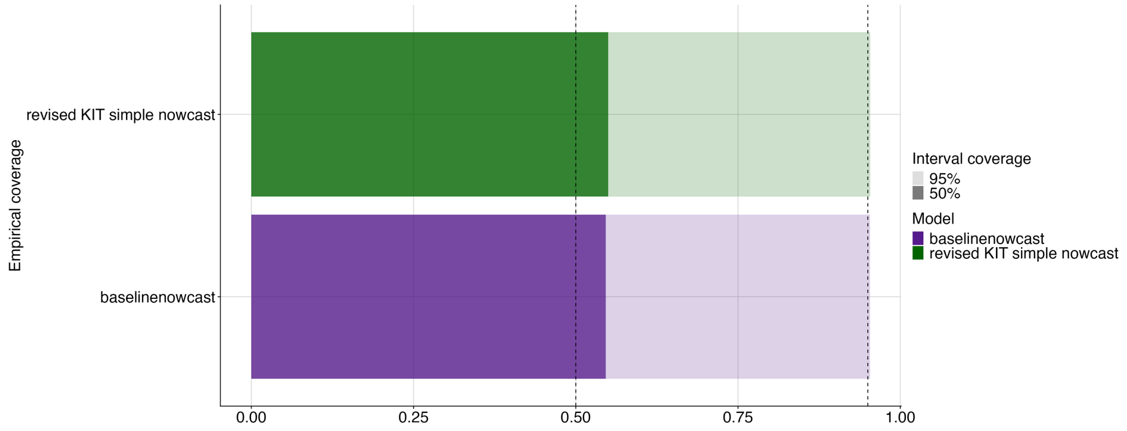


*Fig. S6 Empirical coverage at 50% and 95% prediction intervals. Shading indicates prediction intervals, colour indicates models, vertical lines indicate the target prediction intervals.*

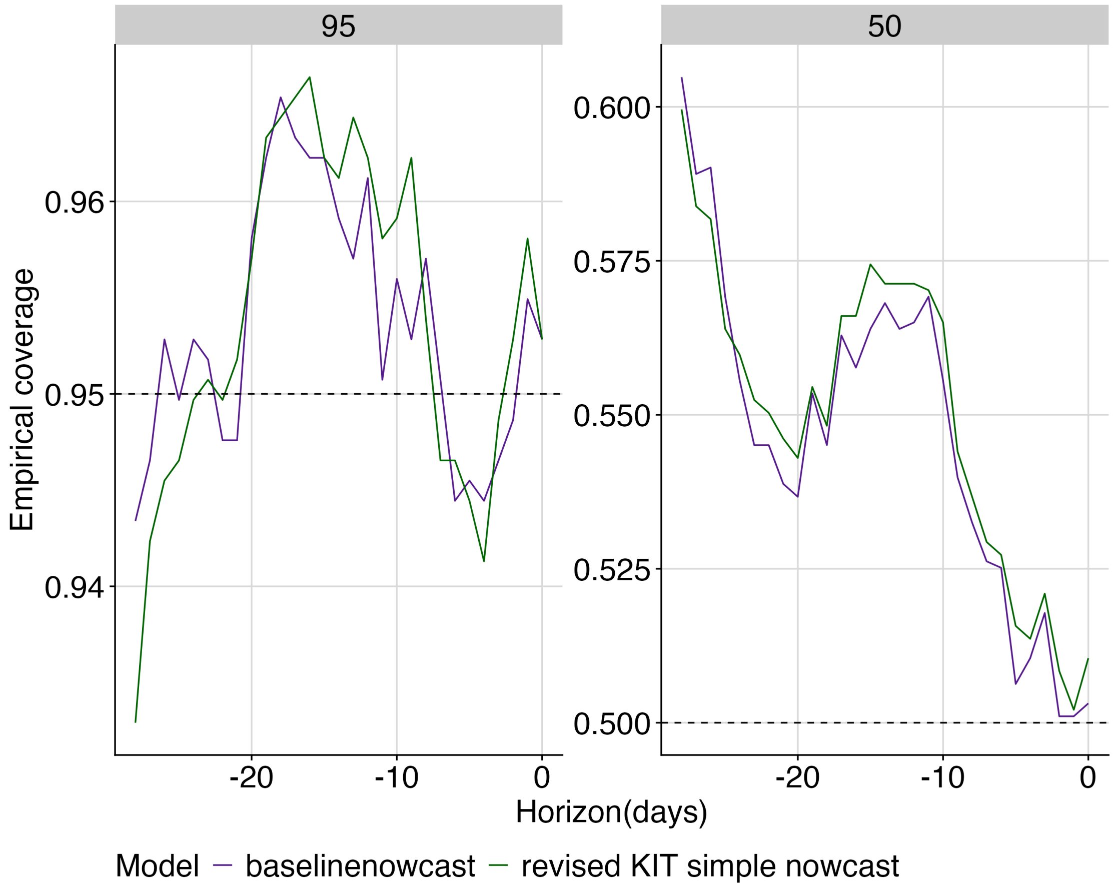


*Fig.S7 Empirical coverage at 50% and 95% prediction intervals by horizon for each model. colours indicate models, horizontal lines indicate the target prediction intervals in each subplot.*

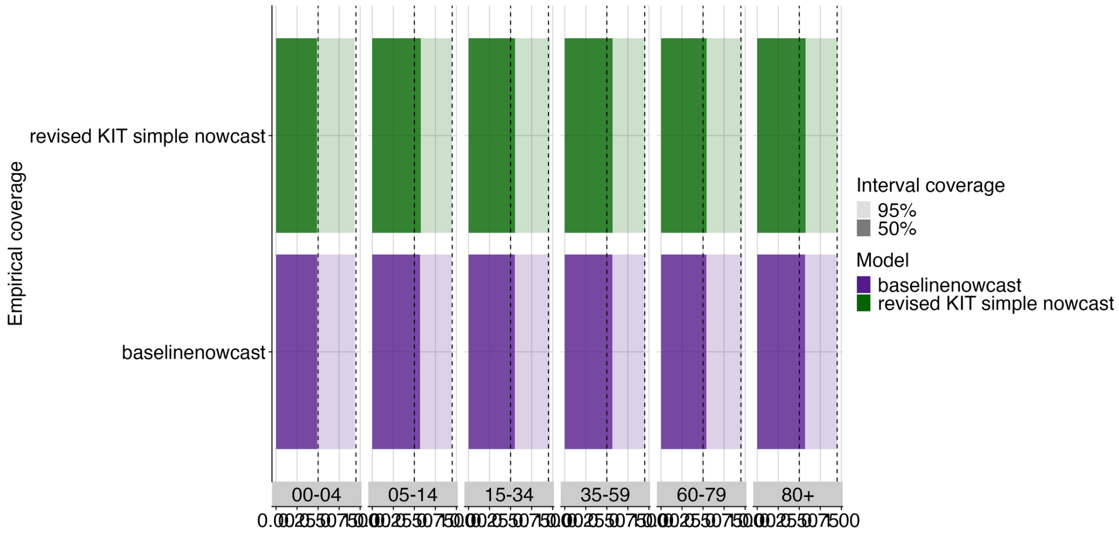


*Fig S8. Empirical coverage at 50% and 95% prediction intervals by age group for each model. Shading indicates prediction intervals, colour indicates models, vertical lines indicate the target prediction intervals.*

#### Performance of different baselinenowcast model specifications applied to German COVID-19 data

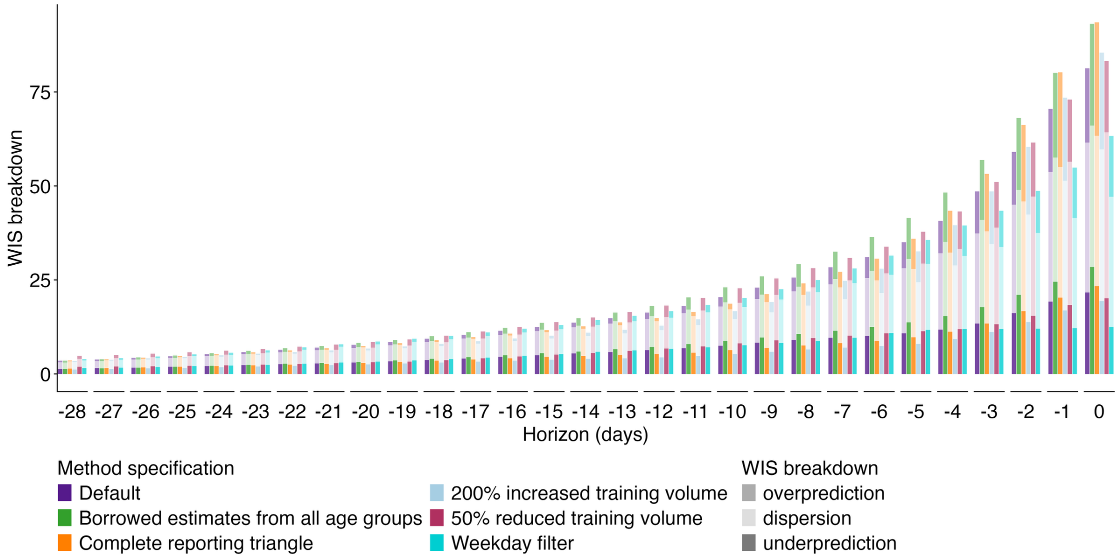


*Fig S9. Absolute WIS by nowcast horizon for each model configuration. Colour indicates model configuration, shading indicates WIS decomposed by dispersion, overprediction, and underprediction.*

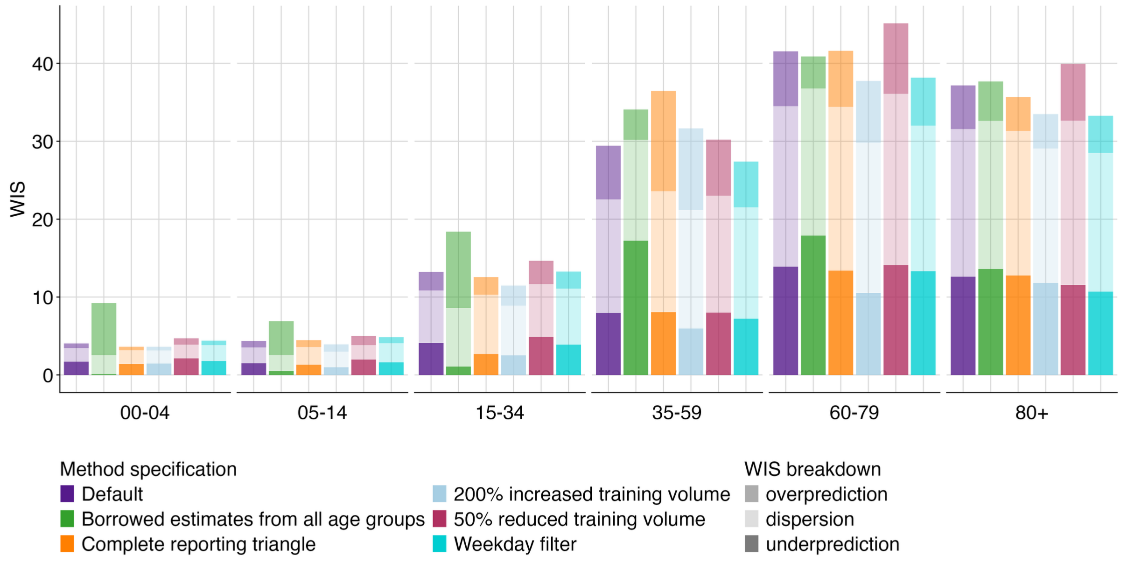


*Fig S10. Absolute WIS by age group for each model configuration. Colour indicates model configuration, shading indicates WIS decomposed by dispersion, overprediction, and underprediction.*

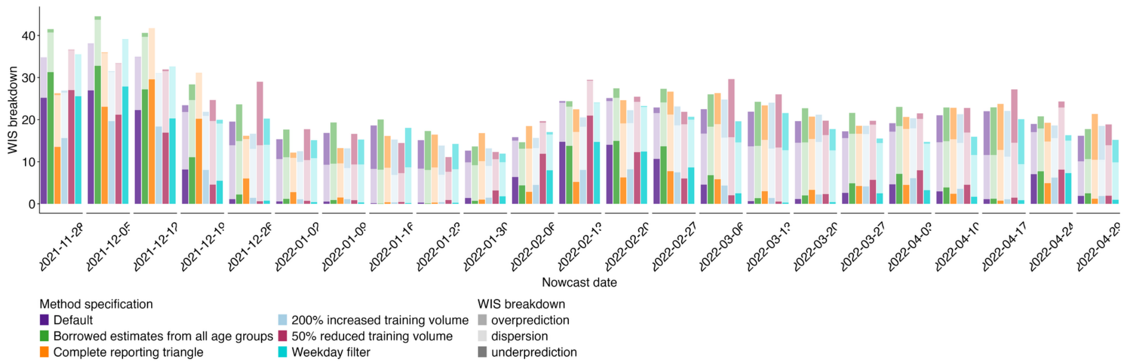


*Fig S11. Absolute WIS over time (by week) for each model configuration. Colour indicates model configuration, shading indicates WIS decomposed by dispersion, overprediction, and underprediction.*

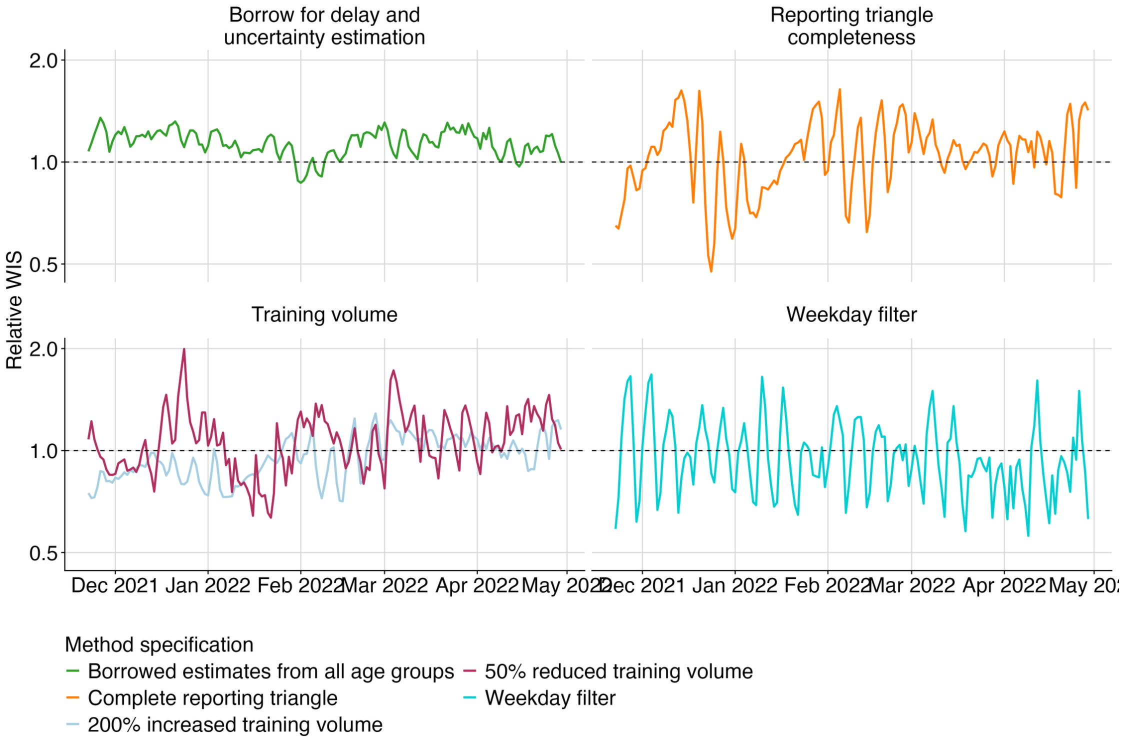


*Fig S12. Relative WIS over time across all age groups for each model configuration. Colours indicate model configuration. Horizontal dashed line indicates parity with the default configuration.*

#### Case study: UKHSA norovirus surveillance

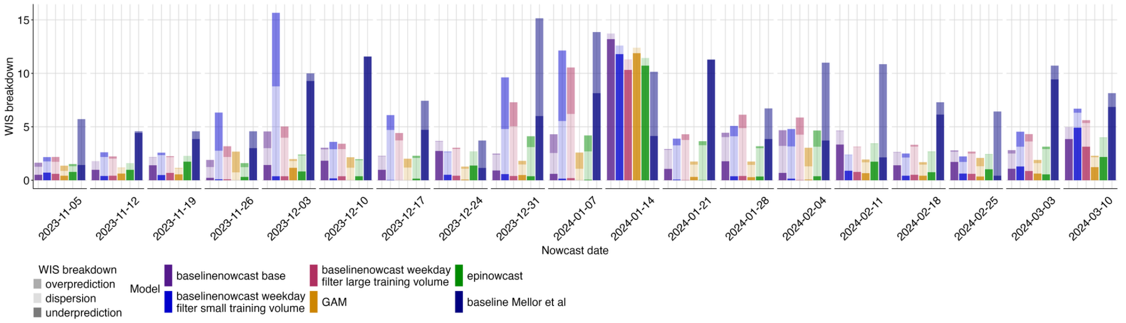


*Fig S13. Absolute WIS over time (by week) for each model. colour indicates model, shading indicates WIS broken down by dispersion, overprediction, and underprediction.*

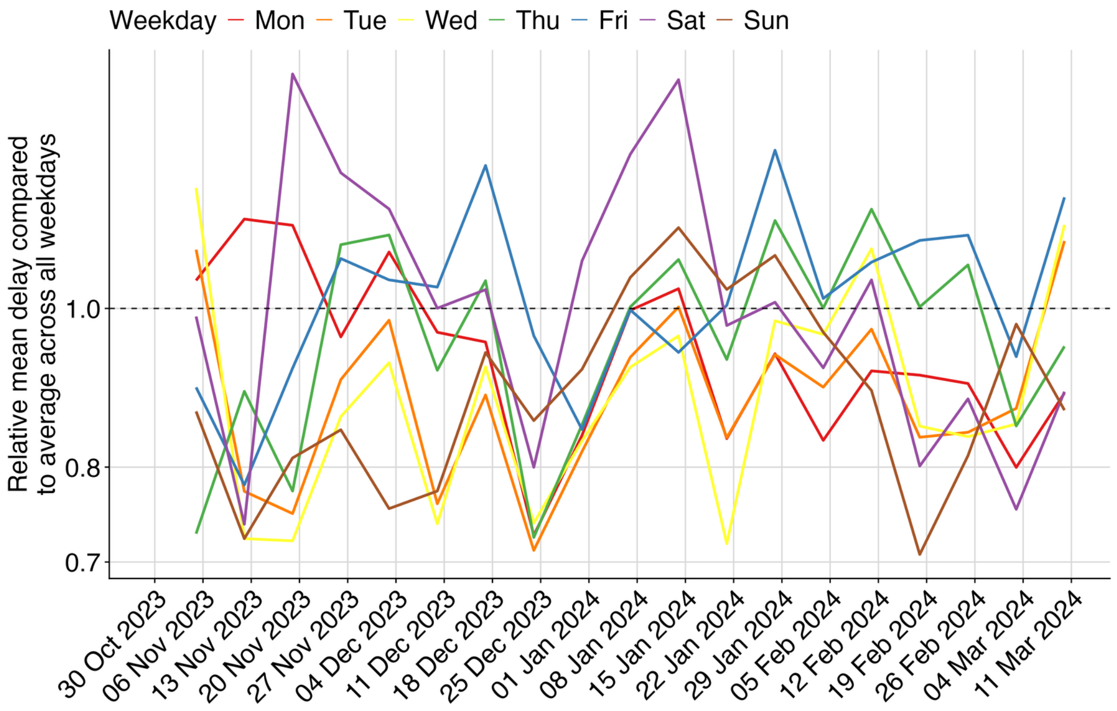


*Fig S14. Relative mean delay over time by weekday. colours indicate weekday. Horizontal dashed line at 1 indicates parity with the delay across all weekdays.*

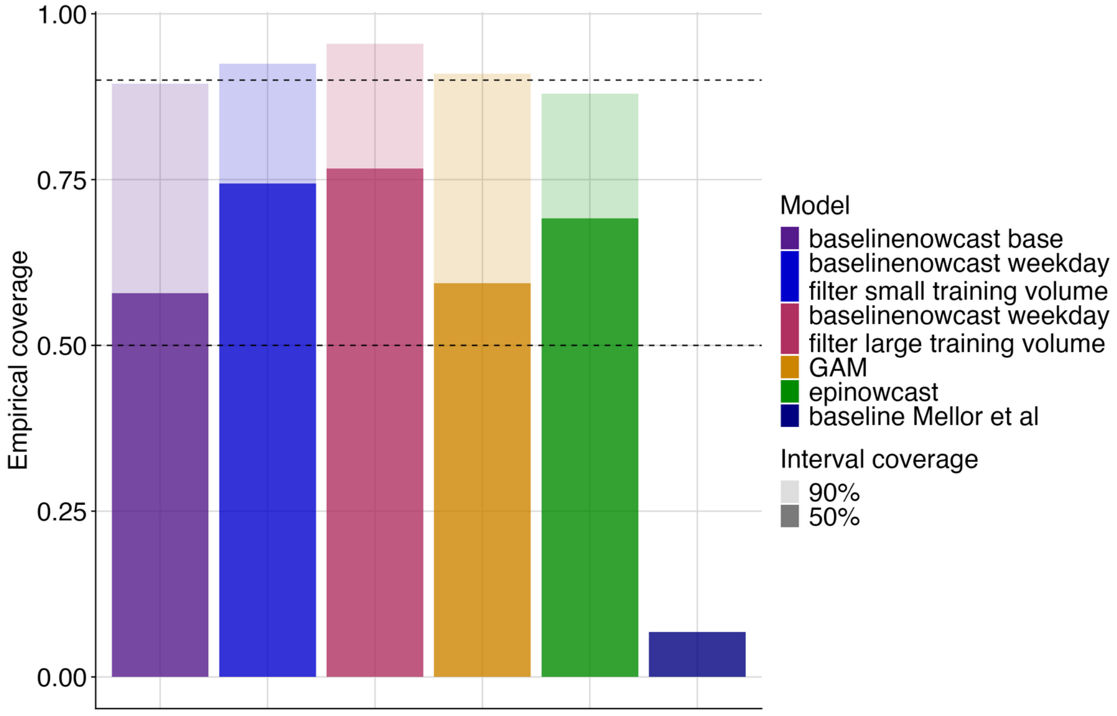


*Fig S15. Empirical coverage at 50% and 90% prediction intervals for each model. colour indicates model, shading indicates prediction interval coverage, horizontal dashed lines indicate target prediction intervals.*

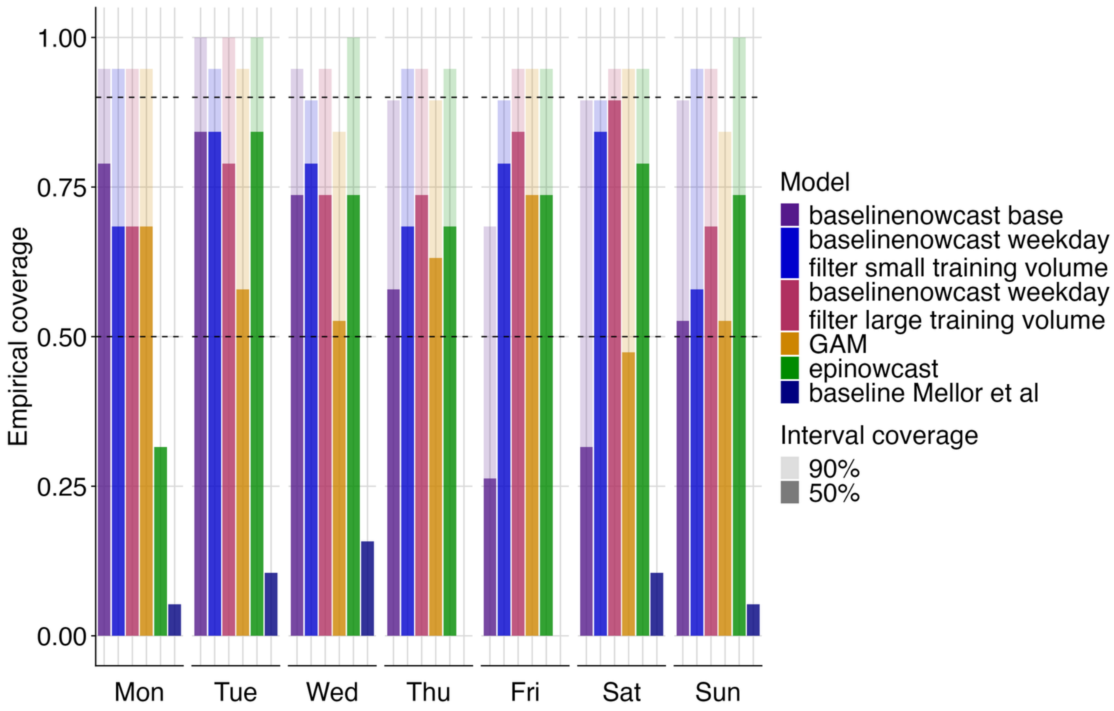


*Fig S16. Empirical coverage at 50% and 90% prediction intervals by day of week for each model. colours indicate model, shading indicates prediction interval coverage, horizontal dashed lines indicate target prediction intervals.*

4. Inc. WR. Mathematica, Version 14.2 [Internet]. Available from: https://www.wolfram.com/mathematica

5. KIT Metrics Lab, Johannes Bracher, Melanie Schienle. RESPINOW-Hub: Nowcasting and Short-term Forecasting Platform for Respiratory Diseases [Internet]. Karlsruhe Institute of Technology; GitHub; 2025 [cited 2025 Jun 25]. Available from: https://github.com/KITmetricslab/RESPINOW-Hub

6. KIT Metrics Lab, Johannes Bracher, Daniel Wolffram, Melanie Schienle. Hospitalization Nowcast Hub: Collecting nowcasts of the 7-day hospitalization incidence in germany [Internet]. Karlsruhe Institute of Technology; GitHub; 2025 [cited 2025 Jun 25]. Available from: https://github.com/KITmetricslab/hospitalization-nowcast-hub
